# fMRI and Transcranial Electrical Stimulation (tES): A systematic review of parameter space and outcomes

**DOI:** 10.1101/2020.06.03.20121202

**Authors:** Peyman Ghobadi-Azbari, Asif Jamil, Fatemeh Yavari, Zeinab Esmaeilpour, Nastaran Malmir, Rasoul Mahdavifar-Khayati, Ghazaleh Soleimani, Yoon-Hee Cha, A. Duke Shereen, Michael A. Nitsche, Marom Bikson, Hamed Ekhtiari

## Abstract

The combination of non-invasive brain stimulation interventions with human brain mapping methods have supported research beyond correlational associations between brain activity and behavior. Functional MRI (fMRI) partnered with transcranial electrical stimulation (tES) methods, i.e., transcranial direct current (tDCS), transcranial alternating current (tACS), and transcranial random noise (tRNS) stimulation, explore the neuromodulatory effects of tES in the targeted brain regions and their interconnected networks and provide opportunities for individualized interventions. Advances in the field of tES-fMRI can be hampered by the methodological variability between studies that confounds comparability/replicability. In order to explore variability in the tES-fMRI methodological parameter space (MPS), we conducted a systematic review of 222 tES-fMRI experiments (181 tDCS, 39 tACS and 2 tRNS) published before February 1, 2019, and suggested a framework to systematically report main elements of MPS across studies. We have organized main findings in terms of fMRI modulation by tES. tES modulates activation and connectivity beyond the stimulated areas particularly with prefrontal stimulation. There were no two studies with the same MPS to replicate findings. We discuss how to harmonize the MPS to promote replication in future studies.

## 1. Introduction

Interest in the use of non-invasive brain stimulation (NIBS) methods to explore and alter the physiological mechanisms underlying basic cognitive processes has grown tremendously over the last several decades, resulting in innovative treatments for several neurological and psychiatric disorders (Brunoni et al., 2019; Schulz et al., 2013). Transcranial electrical stimulation (tES) has especially gaining widespread adoption in laboratories. tES involves delivery of low intensity direct (tDCS), alternating (tACS), or random noise (tRNS) currents, usually between 1-2 mA in intensity, to the brain through electrodes positioned over the scalp (Woods et al., 2016). Application of low-intensity subthreshold neuronal stimulation—normally resulting in an electric field in the brain below 1 V/m (Datta et al., 2009; Huang et al., 2017)— sets tES apart from other popular NIBS techniques such as transcranial magnetic stimulation (TMS), in which stronger electromagnetic pulses (~70-140 V/m induced electric fields (Deng et al., 2013)) are applied (Barker et al., 1985). When tES is applied for several minutes, it can generate neuromodulation effects not only during stimulation, but outlasting stimulation (Bachinger et al., 2017; Nitsche and Paulus, 2001, 2000). The immediate and long-lasting effects of tES, its safety and tolerability (A. Antal et al., 2017; Bikson et al., 2016), non-complex technical requirements, and low cost (Woods et al., 2016) have made tES an attractive option for studying and modulating cognitive, motor, and behavioral processes.

The number of studies that have investigated the use of tES has surged over the past two decades (A Antal et al., 2017; Bikson et al., 2016). This has resulted in numerous systematic reviews and meta-analyses of the role of tES on enhancing cognitive function (Simonsmeier et al., 2018), motor learning (Buch et al., 2017), and clinical applications (Lefaucheur et al., 2016). The wide range of methodological variation of these studies has raised the question of how tES can be used most effectively (Bikson et al., 2018). With increased application, the need for a comprehensive understanding of tES physiological mechanism of action is critical.

A considerable number of studies devoted to this effort have used human, animal, cellular, and computational models to produce evidence for the acute effect of direct (DC) or alternating current (AC) at the single neuron level (Barbati et al., 2019; Cancel et al., 2018; Jackson et al., 2016; Mishima et al., 2019) up to the network level (Monai and Hirase, 2016; Reato et al., 2013), as well as showing lasting after-effects observed in structurally or functionally connected networks (Esmaeilpour et al., 2019; Kuo et al., 2016; Stagg et al., 2018). From these studies, the basic physiological mechanisms of tES on local and regional neuronal activity have become better understood, though the majority of evidence has been observed in the motor cortex. Seminal studies in humans used pharmacological interventions combined with transcranial magnetic stimulation (TMS) as a probe for measuring cortical excitability in the primary motor cortex (M1). These studies found that stimulation after-effects depend on NMDA receptor and GABA activity (Liebetanz et al., 2002; Nitsche et al., 2004, 2003; Stagg and Nitsche, 2011), which form the basis of long term potentiation (LTP) or long term depression (LTD)-mediated learning processes. However, since the number of physiological studies on non-motor areas has been relatively sparse, the generalizability of motor findings to non-motor areas has been limited. Moreover, it has become increasingly evident that the effects of tES are not spatially restricted to the region directly underneath the electrode for reasons of both distributed current flow (Datta et al., 2009) and brain connectivity (Abellaneda-Pérez et al., 2019; Mondino et al., 2019); tES affects multiple, anatomically distant but functionally connected regions (Chib et al., 2013; Stagg et al., 2013; Violante et al., 2017).

A step towards unwrapping the mechanism of tES occurred with the advent of MR-compatible tES devices that made concurrent tES-fMRI technically feasible. These studies can be conducted without posing severe quality constraints, as long as proper procedures are followed (Antal et al., 2014; Esmaeilpour et al., 2019). The special advantage of combining tES and fMRI with a task paradigm makes it possible to explore and modulate causal brain-behavior interactions. Using the so-called “perturb-and-measure” approach (Paus, 2005), the causal/functional contributions of a stimulated brain region to cognitive and perceptual processing can be assessed to answer basic questions about underlying physiology.

A second motivation for combining fMRI with tES has been to determine predictive biomarkers to identify people who are responsive to tES (Esmaeilpour et al., 2019). This can take the form of antatomical and/or functional scans, which can be collected before and/or after tES. Interindividual variability in response to tES has been reported across many studies (Chew et al., 2015; Kim et al., 2014; Li et al., 2015), and can be explained by both anatomical differences impacting current flow (Datta et al., 2012, 2011; Edwards et al., 2013; Evans et al., 2020; Gomez-Tames et al., 2020) and brain activity impacting brain-state specific outcomes (Bikson et al., 2013; Li et al., 2019; Nishida et al., 2019). Obtaining structural and functional MRI information prior to tES allows determination of factors such as gray and white matter structure or functional activation or connectivity to predict response to certain tES interventions (Cavaliere et al., 2016; Datta et al., 2012; Hordacre et al., 2017; Laakso et al., 2015; Opitz et al., 2015). Thirdly, tES-fMRI aids in the optimization of electrode montages using fMRI data integrated with anatomical and physiological parameters such as tissue conductivity, cortical anatomy (including thickness and volume), and blood perfusion patterns (Baker et al., 2010; Fischer et al., 2017). These parameters can aid in the development of individualized stimulation montages and protocols based on individual or group level functional organization of the targeted cognitive function. These fMRI informed montages might also be helpful for clinical applications such as in stroke in which the lesion influences both functional organization and the pattern of the induced electric field (Datta et al., 2011; Wagner et al., 2007). The three aims of tES-fMRI studies are schematically summarized in Figure 1.

**Figure 1:**
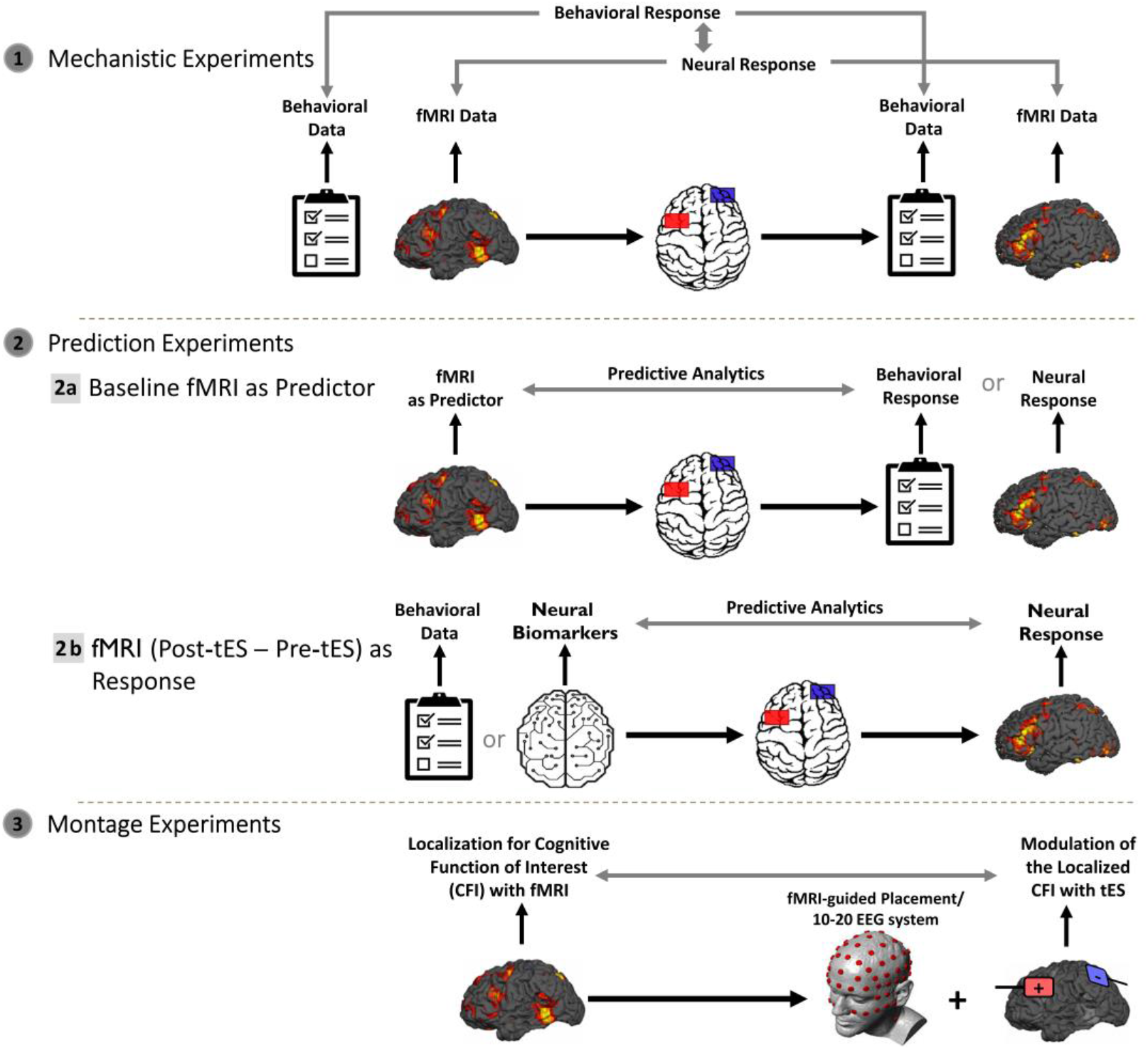
Three categories of tES-fMRI studies based on the role of fMRI. (1) Mechanistic experiments, which use fMRI to investigate the mechanisms underlying tES effects. The ultimate aim of these experiments is to find a relationship between behavioral and neural responses to tES**. (2)** Predictive experiments: **(2a)** Investigate how baseline fMRI measures can predict behavioral/neural responses to tES (i.e., “Baseline fMRI activity as Predictor”); **(2b)** Use behavioral or neural biomarkers like structural neuroimaging data to predict the neural responses to tES as measured with fMRI (i.e., “fMRI (Post-tES-Pre-tES) as Response”). **(3)** Montage experiments, in which fMRI is used to localize the brain areas whose activation correlate with certain cognitive functions of interest. tES is then used to modulate these areas in order to elucidate their causal role in these functions. Details and subcategories of each of the three main categories can be found in Figures 3, 7 and Table 1.

Inherent in the design of any fMRI study, and more critical when combined with tES, is the consideration of how methodological parameters impact interpretation of findings and which methodological parameters are important to control and consider when running and contrasting experiments. When reviewing the published studies, we noted the reporting of a wide range of practices in designing and reporting methodological parameters, spanning both fMRI and stimulation protocols. These parameters shape the “methodological parameter space (MPS)” for tES-fMRI studies. These parameters include categorical factors such as: the specific electrode montage, equipment selection and configuration, inclusion of a sham condition, and continuous factors such as the tES intensity, duration, number of sessions, physiological, and behavioral outcomes. There are additional factors such as the fMRI scanning sequence and whether scans were acquired pre-, online, or post-tES. These factors interact in a combinatorial manner creating considerable heterogeneity, which precludes accurate groupings of even seemingly similar studies.

Following our previous review paper on the methodological considerations in the integration of tES and fMRI (Esmaeilpour et al., 2019), in this systematic review we aimed to consolidate the extant knowledge of the published tES-fMRI studies, where methodology has progressed largely independently between different research groups, resulting in diverse protocols and findings. As such, we have deemed that such a review would be useful for the following reasons: 1) Obtaining an overview of the extant tES-fMRI research and its MPS which would be helpful for detecting trends and gaps in the field; 2) In order to advance the mechanistic knowledge of tES effects, a meta-analysis on fMRI findings in tES-fMRI studies is needed to determine how this intervention works; 3) Mapping the inconsistency of the methodological approaches in tES-fMRI studies will be critical to understand the heterogeneous mixture of findings, which cannot always be interpreted independently from the methodological parameters (Nitsche et al., 2015); 4) Given the large and increasing number of research labs using tES-fMRI, this systematic review provides a methodological framework to categorize and harmonize MPS in future tES-fMRI studies.

## 2. Methods

This systematic review was conducted in accordance with the most updated Preferred Reporting Items for Systematic Reviews and Meta-Analyses (PRISMA) guidelines (Moher et al., 2010).

### 2.1. Search Strategy

Two independent reviewers (PGA and NM) conducted a literature search for relevant studies utilizing the PubMed research database from inception up to February 1, 2019. The search terms were a logical combination of keywords (“tDCS” OR “transcranial direct current stimulation” OR “tACS” OR “transcranial alternating current stimulation” OR “tRNS” OR “transcranial random noise stimulation”) AND (“functional magnetic resonance imaging” OR “fMRI” OR “functional MRI” OR “fcMRI” OR “functional connectivity MRI” OR “rs-fMRI” OR “resting-state fMRI”).

### 2.2. Study Selection Process

Studies had to report the fMRI technique (BOLD or ASL) with its paradigms and how fMRI was combined with tES (including tDCS and tACS) as well as to contain original results. Books, case reports, abstract-only articles, guideline articles, protocol and review articles, studies which combined fMRI with brain stimulation techniques other than tES (e.g., DBS, ECT, and TMS), studies in which tES was only combined with neuroimaging measures other than fMRI (e.g., diffusion tensor imaging (DTI)/diffusion-weighted imaging, magnetic resonance spectroscopy (MRS), and structural MRI), and/or studies which combined tES only with electrophysiological techniques (e.g., EEG, MEG, and event-related potentials) were excluded. tRNS-fMRI studies were not considered in this systematic review, because only two respective studies were published so far (Chaieb et al., 2009; Saiote et al., 2013). The list of all included studies was explored by independent investigators, limiting the search to studies published in English and trials performed on human subjects (healthy or patient subjects).

### 2.3. Extracted Variables

Two investigators independently screened the titles and abstracts of the 493 full-length publications without reviewing any data and excluded 239 studies based on the predefined exclusion criteria to yield 254 studies (Figure 2. PRISMA flow chart). Subsequently, the full text of each of the 254 studies was carefully evaluated, resulting in further exclusion of 116 studies, with specific reasons given in the PRISMA flow chart (Figure 2). This yielded the final 138 studies included in the qualitative synthesis. In those studies that collected data in more than one experimental condition (different montages, polarities, frequencies, stimulation intensities and stimulation durations), each experimental condition was treated as a unique experiment. For example, in one study, Antonenko et. al., compared the effects of 1mA DC stimulation for 15 minutes using C3/Fp2 and Fp2/C3 montages with sham (Antonenko et al., 2017). We treated each active stimulation condition (anodal or cathodal over C3) relative to the sham condition (active vs sham) as a individual experiment, and extracted the stimulation/imaging parameters and the outcome for each of these experiments separately. In total, the 138 reviewed studies generated 220 experiments (Figure 2). This systematic review was accomplished independently by the researchers using the same bibliographic search strategy. Any discordance related to a study’s eligibility criteria was subsequently resolved through discussion with the senior researcher (HE).

**Figure 2:**
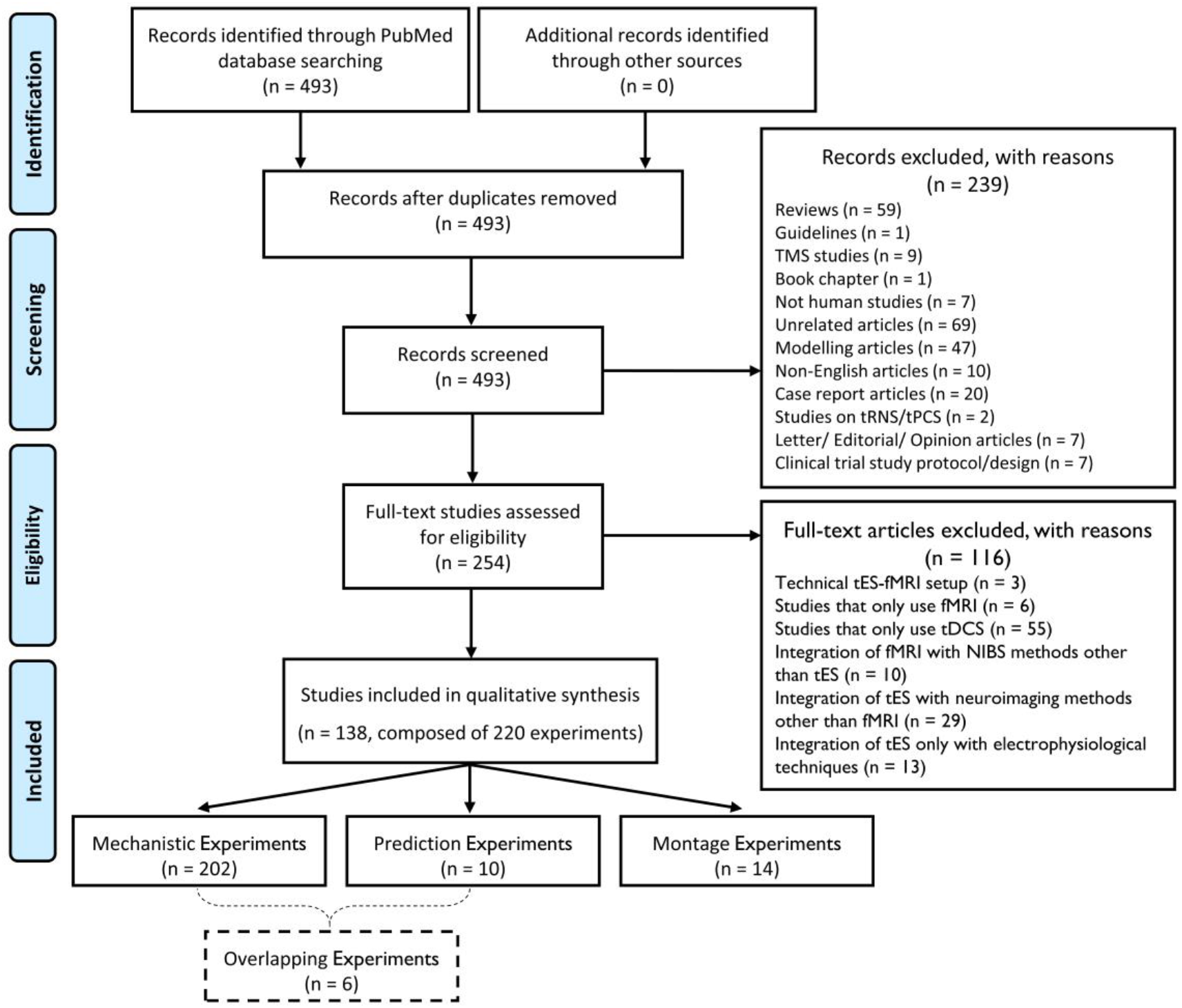
PRISMA flowchart for the selection of studies. In those studies that collected data in more than one experimental condition (different montages, polarities, frequencies, stimulation intensities, and stimulation durations), each experimental condition was treated as a unique “experiment”. Therefore, the 138 reviewed studies included a total of 220 experiments.

## 3. Results

### 3.1. Main Categories of Experiments

One hundred thirty-eight studies were published between January 1, 2000 and February 1, 2019 that used tES in combination with fMRI. Based on the role of fMRI, these studies were classified into three main categories (Figure 1): (1) mechanistic experiments (n=202); (2) predictive experiments (n=10, 6 of them also categorized as mechanistic); and (3) montage experiments (n=14). In the following sections, each of the three categories and the main subcategories in each are described in more detail.

### 3.2. Mechanistic Experiments

Transcranial electrical stimulation can be combined with fMRI to reveal network level changes induced by stimulation. Among the identified experiments, mechanistic experiments (i.e., experiments specifically aimed at fMRI signal alterations induced by tES) constitute the majority, with 202 experiments (163 tDCS, 39 tACS). The MPS in these experiments is summarized in Figure 3. These experiments used different combinations of the following research questions: 1) understanding the impact of tES on the fMRI signal as a proxy measure for local and global neuro-metabolic activity, 2) exploring the behavioral (including selfreported) outcomes of tES (independent to the fMRI) and 3) determining the relationship between the effect of tES on fMRI signal and behavior, if any.

**Figure 3:**
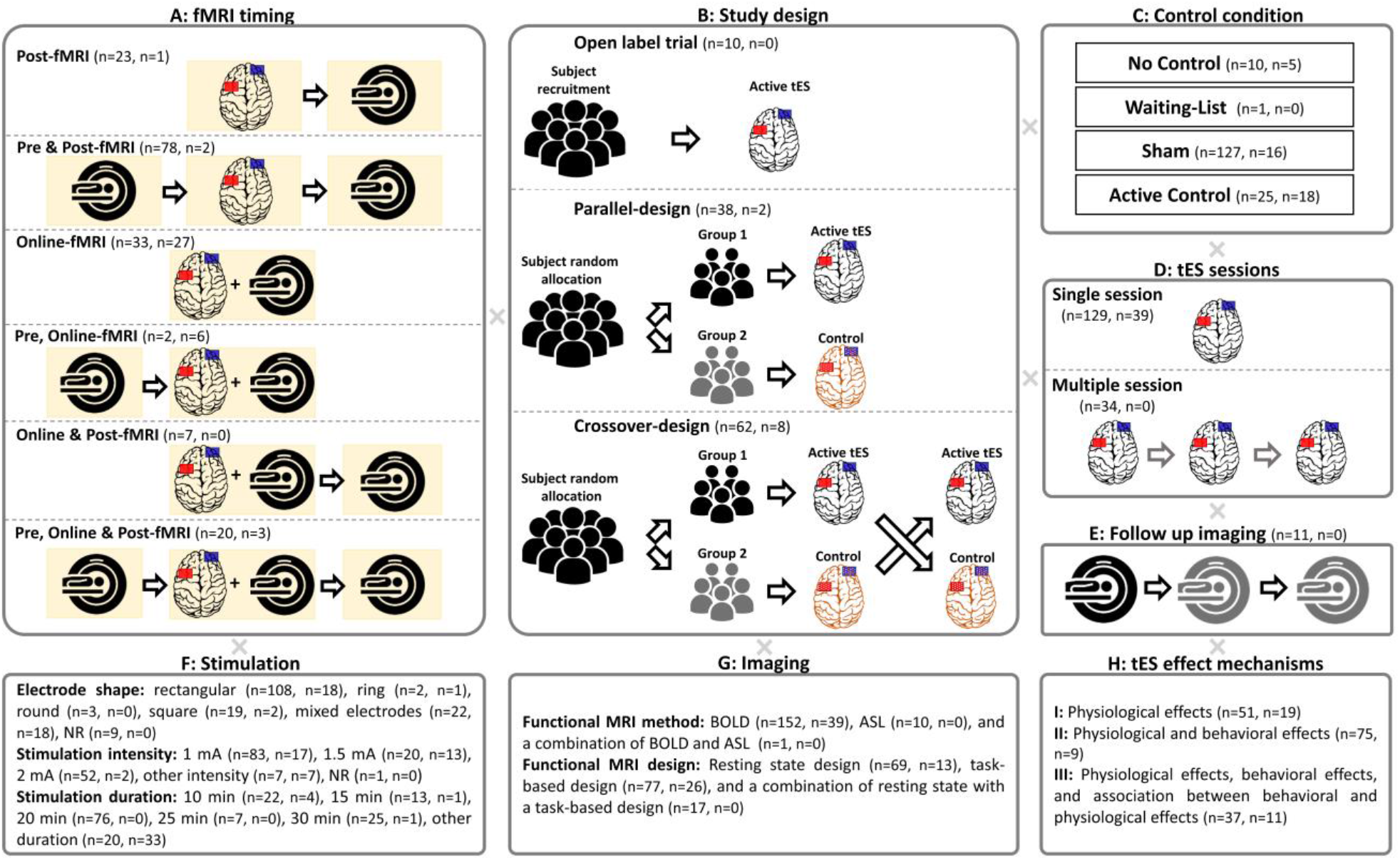
Factorial nature of the methodological parameter space for tES mechanistic experiments (163 tDCS, 39 tACS) using fMRI. **(A)** fMRI was performed before, during, or after stimulation. **(B)** Study designs were open label, parallel, or within-subject crossover. **(C)** Control conditions were active control and/or sham tES. **(D)** tES could be applied in multiple sessions in order to observe cumulative effects. **(E)** Multiple follow-up imaging sessions could be employed. **(F)** Stimulation parameters (i.e., electrode shape, stimulation duration and intensity) can be investigated. **(G)** Functional imaging sequences varied depending on the objective of the study. **(H)** Different mechanisms of tES effect were investigated which include different combinations of physiological effects, behavioral effects, and the association between physiological effects with behavioral findings. Numbers in parentheses represent the number of tDCS and tACS experiments, respectively. Note in the study design panel, the reported numbers refer to the number of studies, not experiments.

#### 3.2.1 Mechanistic tDCS/fMRI Experiments

When organized by experimental design and analysis pathways, 32% of all tDCS/fMRI mechanistic experiments (n=52) investigated physiological effects of tDCS (step 1), 46% (n=75) reported effects of tDCS on both behavior and physiology (including both steps 1 and 2 independently), and the remaining 22% (n=36) took all 3 steps and reported associations between behavioral and physiological effects of tDCS (steps 1, 2 and 3) (Figure 3H).

We further categorized these experiments by the *target* region of neuromodulation, based on the electrode which was placed over the putative target region (and not a control region). This categorization revealed a prevailing trend whereby more than half of the experiments either targeted the sensorimotor cortex (44% of all montages, n=71), or the prefrontal cortex (PFC) (36%, n=58) (see Figure 4 for overview of the categorization). Moreover, we categorized the reported fMRI results across all experiments in order to ask whether stimulation to a given targeted area showed physiological effects confined to the targeted area or whether effects were also observed outside of the targeted area. As shown in Figure 5, mechanistic findings were not necessarily constrained to brain areas directly targeted by the stimulation montage, rather widespread effects were observed across a number of experiments. Strikingly, the most widespread effects were observed with experiments which targeted the PFC, showing that areas across the entire brain were affected, including the parietal cortex, as well as subcortical areas. We also observed considerable heterogeneity in experimental methodology, related to the choice of tES montage, current intensity, duration, etc., and also with fMRI features, such as imaging sequence, scanner timing, and paradigm design. As such, we further categorized experiments according to these fMRI and tES methodological parameters.

**Figure 4:**
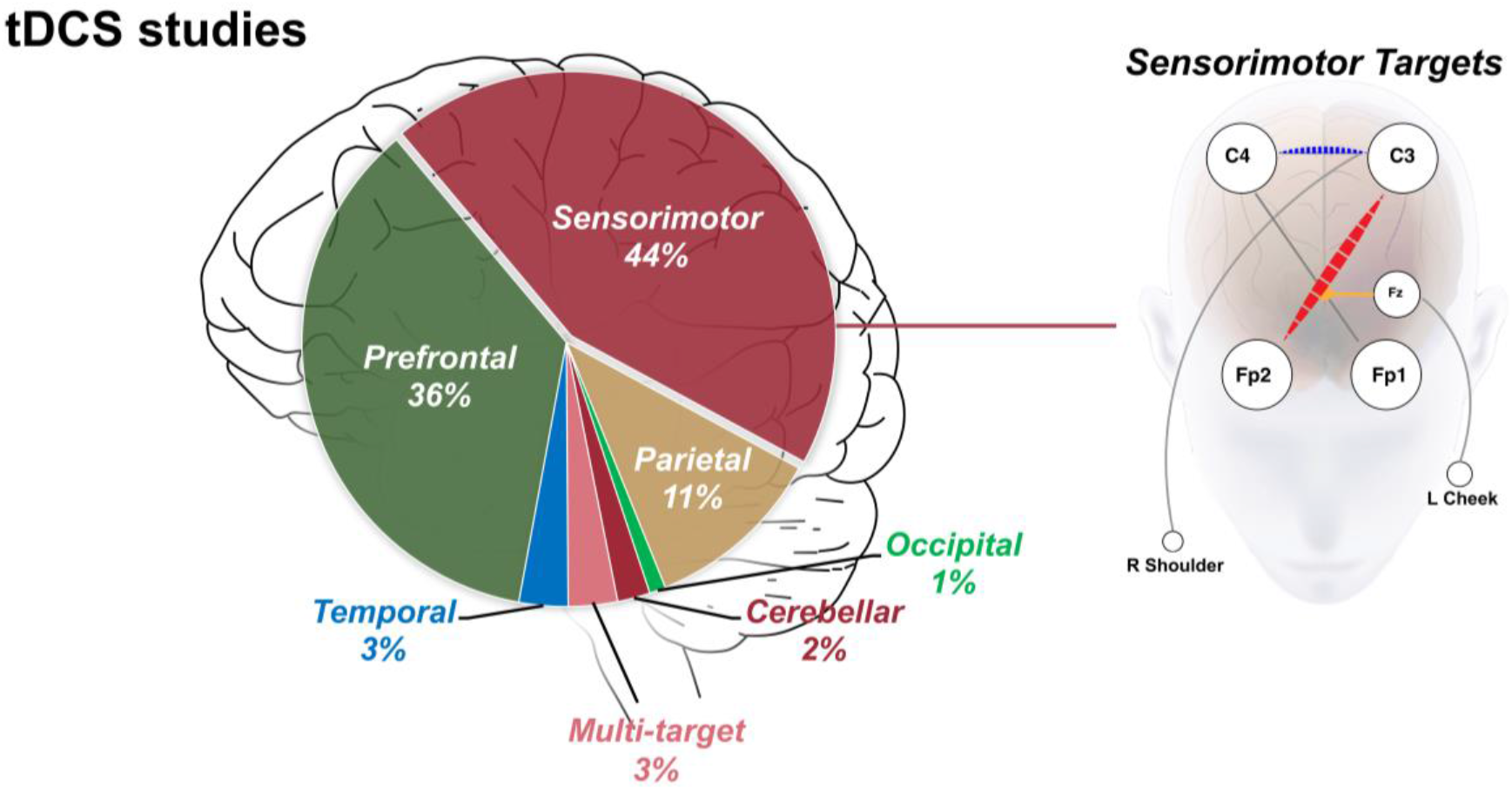
Relative distribution of stimulation target areas across tDCS mechanistic experiments. Details of electrode montages were extracted to identify the major target area of stimulation. As can be seen, the largest portion of tES-fMRI mechanistic experiments target the sensorimotor area (44%), followed by prefrontal (36%), and parietal area (11%). On the right side, the variety of montages targeting the sensorimotor area are depicted by circles and lines. The red line connects the location of the electrodes in the most frequently used montage, and the blue line for the second most frequently used.

**Figure 5:**
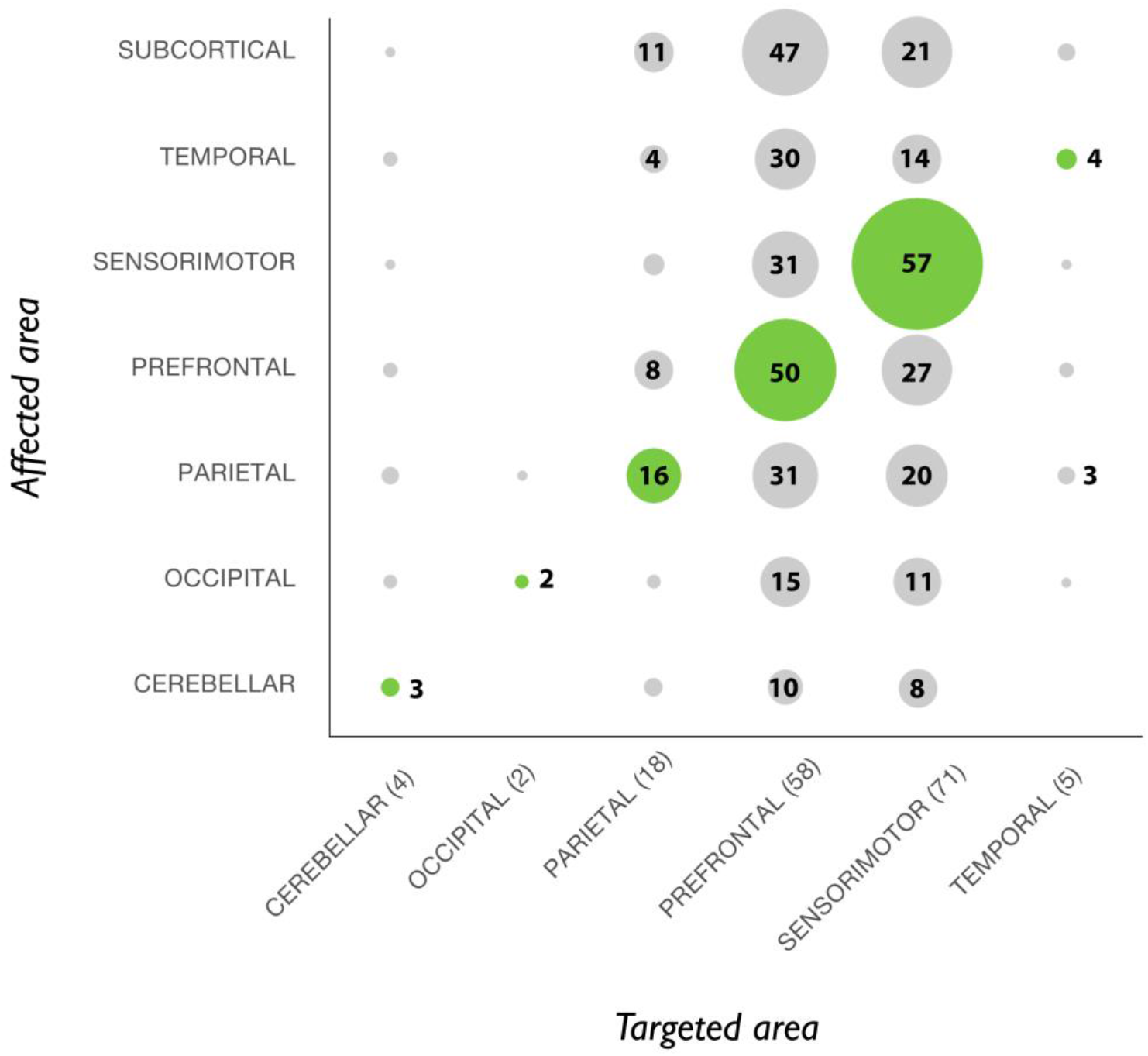
Mapping stimulation targets with physiological findings. For each identified mechanistic experiment, we asked whether the observed target area of stimulation resulted in the largest effect within the target area, or elsewhere. The horizontal axis shows the target areas that were defined based on the position of the electrodes (with the total number of experiments within the respective target area specified in the parentheses). The vertical axis shows the affected areas. The size of each circle is proportional to the number of the corresponding experiments, with the green color marking the diagonal. The majority of evidence shows that targeting the sensorimotor areas have maximum effect on these areas but that stimulation effects can be widespread. Of note, some experiments have targeted more than one area.

With regard to fMRI parameters, 42% (n=69 experiments) used a resting-state design, and the remaining used either a task-based (47%, n=77) or a combination of resting-state with a task-based design (10%, n=17). The majority of experiments used the blood oxygen level–dependent (BOLD) sequence (93%, n=152) compared to arterial spin labeling (ASL) (6%, n=10) (one experiment used both BOLD and ASL techniques). The most common stimulation-imaging timing used was pre-post (49%, n=78 experiments), followed by online (20%, n=33), and post stimulation designs (14%, n=23). Most of the experiments enrolled healthy participants (young or elderly, 76%, n=124), whereas the remainder included populations of different patient groups. With regard to the electrode montage, among those experiments which reported the electrode shape for the main stimulation electrode (95%), most experiments used pairs of rectangular electrodes (66%, n=108), as compared to square electrodes (12%, n=19), round electrodes (2%, n=3), or ring shape electrodes (1%, n=2). Different electrode sizes were used, with a majority however using 35 cm^2^ (69%, n=112). The current intensity applied in these experiments ranges from 0.5 to 2 mA, with a majority of experiments using 1 mA (51%, n=83). The total duration of the stimulation ranges from 0.33 to 30 minutes, with a majority of experiments stimulating for 20 min (47%, n=76). The positioning of electrodes on the scalp, in those experiments which reported the respective method, were determined either using physiological markers (18%, n=30) (e.g., TMS MEP hotspot or fMRI activation), neuro-navigation (4%, n=7) or the 10-20/10-10/distance-based system (72%, n=117). Relative positioning of the anodal and cathodal electrodes was mostly bilateral bipolar-non balanced (54%, n=88), followed by bilateral bipolar balanced (18%, n=30) (Nasseri et al., 2015). From the 110 studies, 62 used a crossover design, 38 were performed with a parallel design, and 10 studies were open label trials. With regard to the control condition in each experiment, 127 used sham stimulation, 25 had an active control, 10 were performed without any control condition, and only one experiment used a “waiting-list as control” design. Twenty-two experiments employed an active control strategy, either using a different stimulation site, but same polarity, or the same stimulation site with a different polarity.

#### 3.2.2 Mechanistic tACS/fMRI Experiments

When organized by experimental design and analysis pathways, 49% of all tACS mechanistic experiments (n=19) investigated physiological effects of tES (step 1), 23% (n=9) reported effects of tES on both behavior and physiology (including both steps 1 and 2 independently), and the remaining 28% (n=11) took all 3 steps and reported associations between behavioral and physiological effects of tACS (steps 1, 2 and 3). We further categorized these experiments by the *target* region of neuromodulation, based on the location of the electrode that was placed over the putative target region. We observed that over 72% of all tACS experiments targeted the occipital cortex, with most of these using a stimulation frequency of 16 Hz (32%) (see Figure 6 for a full breakdown).

**Figure 6:**
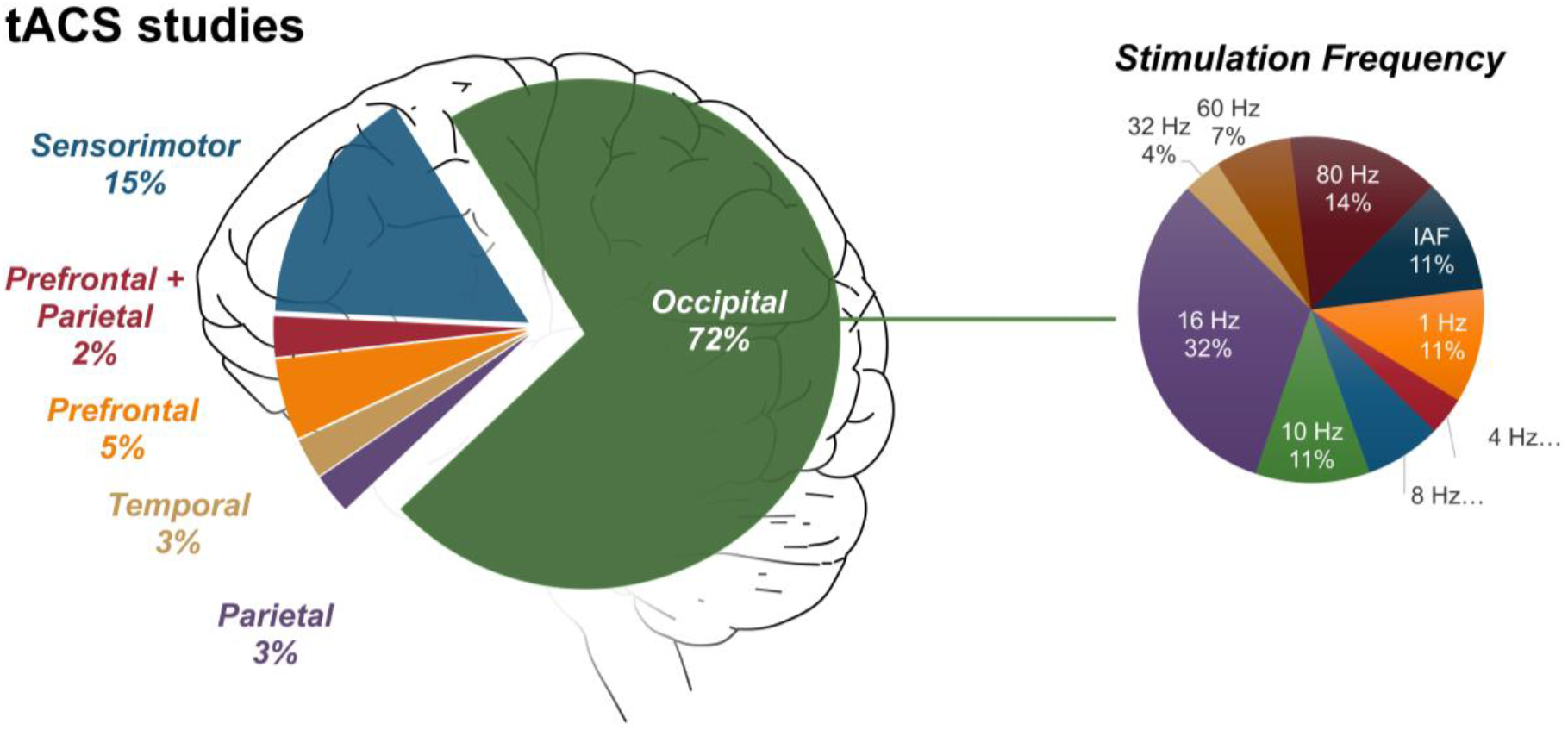
Relative distribution of stimulation target areas across tACS mechanistic experiments identified at present. Details of electrode montages were extracted to identify the major target area of stimulation. As can be seen, the largest portion of tACS-fMRI mechanistic experiments target the occipital area (72%), followed by the sensorimotor (15%), and the prefrontal areas (5%). On the right side, the variety of stimulation frequencies targeting the occipital area are depicted by the pie chart. Most experiments stimulated at a frequency of 16 Hz (32%) followed by 10 Hz (11%).

Similar to the tDCS experiments, we further categorized tACS experiments according to stimulation and previously mentioned fMRI methodological parameters.

With regard to fMRI parameters, 33% (n=13 experiments) used a resting-state design; the remaining used a task-based design (67%, n=26). BOLD-fMRI was used across all fMRI-tACS experiments. The most common stimulation timing was online stimulation design (69%, n=27 experiments), followed by pre vs online stimulation design (15%, n=6), pre-online-post design (8%, n=3 experiments), pre-post design (5%, n=2). Only one experiment had post stimulation fMRI design. All of these experiments enrolled healthy participants.

With regard to the electrode montage, most experiments used pairs of square or rectangular (59%, n=23), as compared to circular or ring electrodes (3%, n=1). The remaining experiments used a mix of round and square electrodes (38%, n=15). Reported electrode sizes range from a majority using 35 cm^2^ for both electrodes (46%, n=18), followed by 16 cm^2^ for one electrode and 35 cm^2^ for the other (38%, n=15). The remaining experiments used more mixed sizes, but these sizes were not applied in more than two experiments in each case. Current intensity ranged from 0.2 to 1.7 mA peak-to-peak, with a majority of experiments having used 1 mA peak-to-peak (49%, n=19). The total stimulation duration ranged from 1.2 to 30 minutes, with a majority of experiments stimulating for 3 (15%, n=6) or 6 min (15%, n=6). The positioning of electrodes on the scalp, in those experiments which reported the method, were most often determined based on the 10-20/10-10/distance-based system (92%, n=36). Relative positioning of tACS electrodes was in most cases midline, bipolar-balanced (77%, n=30). With regard to the control condition in each experiment, 16 used sham stimulation as the control condition, 18 implemented an active control, and 5 were performed without any control condition. Of the 10 available studies, 8 used a crossover design and 2 were performed with a parallel design.

### 3.2. Predictive Experiments

There are 10 published experiments since 2000 which have employed fMRI as predictor/response to predict the neural/behavioral outcomes in response to tES (Figure 7). Depending on using fMRI as predictor or response, these 10 predictive experiments are categorized into two main groups: (1) “fMRI-as-predictor” experiments, which aim to explain the variance in behavioral/neural outcomes in response to tES based on pre-tES fMRI (n=6); and (2) “fMRI-as-response” experiments, which aim to use behavioral or neural biomarkers to predict the neural responses to tES as measured with fMRI (n=4). One study (Polanía et al., 2012) uses fMRI as both, predictor and response, and thus falls into both of the mentioned groups. The following sections provide a more detailed description of these two categories and the MPS for fMRI in each.

**Figure 7:**
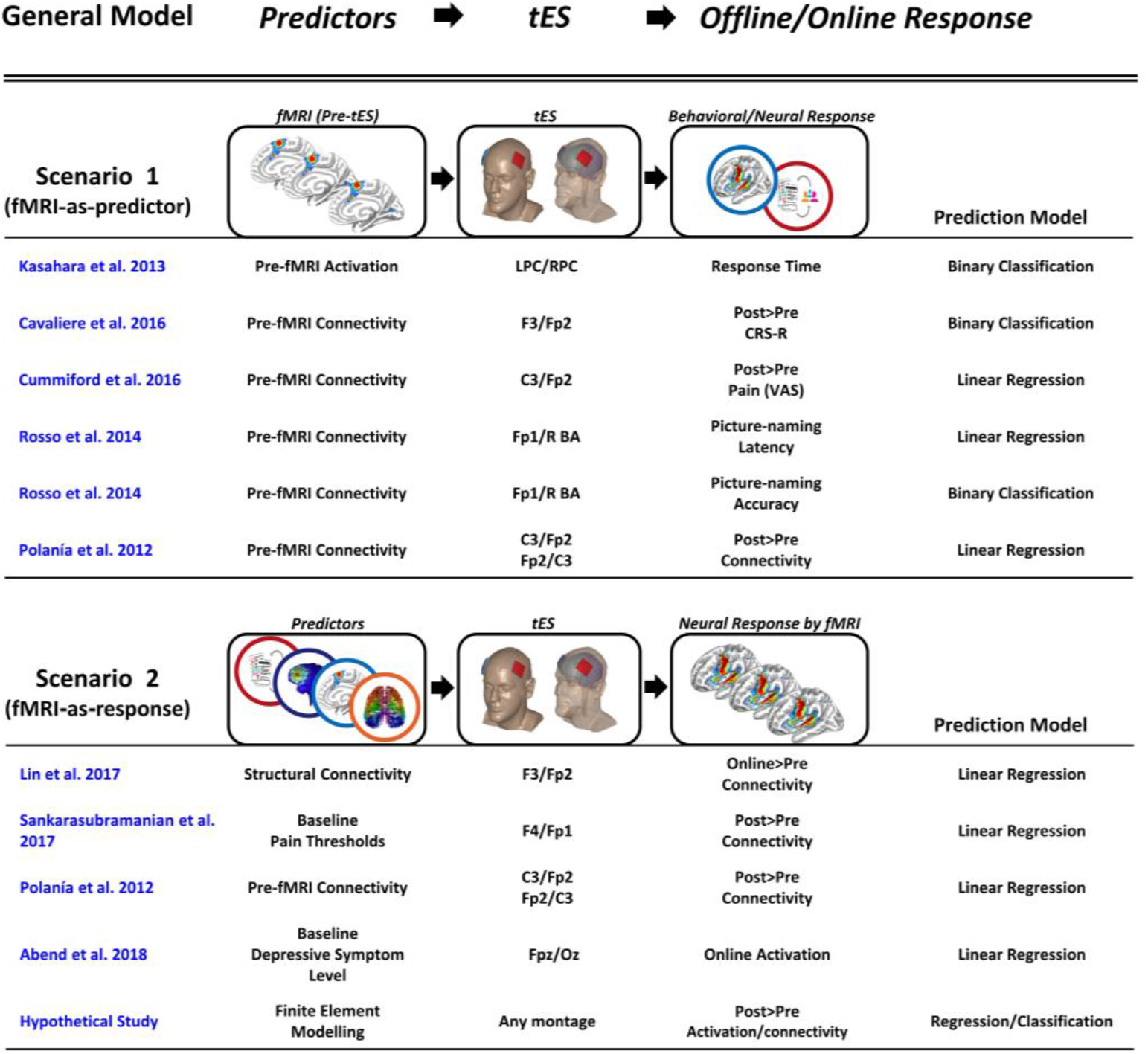
Studies which used fMRI to predict behavioral/neural responses to tES. **Scenario 1**, estimating the prediction power of baseline fMRI for explaining the variance in the behavioral/neural outcomes in response to tES. **Scenario 2**, using fMRI to measure response to tES in predictive modeling. BA: Broca’s area; CRS-R: coma recovery scale revised; LPC: left parietal cortex; R: right; RPC: right parietal cortex; VAS: visual analogue scale.

#### 3.2.1. FMRI-as-Predictor

The “fMRI-as-predictor” approach allows researchers to investigate how pre-tES fMRI measures (e.g., functional activity, or connectivity) predict the subsequent behavioral/neural response to tES (n=6 experiments).

Out of the six “fMRI-as-predictor” experiments, only one experiment used functional activity to predict the behavioral response to tES. Kasahara and colleagues examined the effects of bilateral parietal tDCS on mental calculation task performance in 16 healthy subjects (Kasahara et al., 2013). Baseline lateralization of fMRI activity in the parietal cortex toward the left hemisphere, specifically during mental calculation task performance, correlated with better response to bilateral parietal tDCS. Specifically, in individuals with left-hemispheric dominance of parietal activity, response time was significantly shortened when anodal tDCS was applied to the left parietal cortex and cathodal tDCS was applied to the right parietal cortex. This finding indicates that subjects with left-hemispheric parietal lateralization are most likely to benefit from bilateral parietal tDCS with respect to performance of mental calculations.

Five out of the six “fMRI-as-predictor” experiments have demonstrated a key role of baseline functional connectivity in predicting behavioral/neural responses to tDCS. Cavaliere and colleagues investigated pre-treatment and early treatment rs-fMRI markers of response in 16 minimally consciousness state (MCS) patients undergoing dorsolateral PFC (DLPFC) tDCS (Cavaliere et al., 2016). Six patients were classified as responders, based on post-tDCS minus pre-tDCS Coma Recovery Scale Revised (CRS-R), after a single session tDCS over the left DLPFC (using a F3-Fp2 montage) in a double-blind randomized crossover trial, and ten were non-responders. Higher functional connectivity between left DLPFC and left inferior frontal gyrus (IFG), based on a seed-based rs-fMRI connectivity analysis for regions of the left extrinsic control network (ECN), and default-mode network, at baseline was predictive of behavioral response to tDCS (i.e., post-tDCS minus pre-tDCS CRS-R) in MCS patients. Pre-treatment connectivity between left DLPFC and left IFG was higher in the responders compared to nonresponders. Patients unlikely to respond showed a lower left ECN connectivity with a more diffuse and bilateral co-activation of anterior cingulate cortex and precuneus regions. These findings suggest that a prior high connectivity of ECN regions can facilitate transitory recovery of consciousness in MCS patients that underwent a single session of anodal tDCS.

Response to 5 days of tDCS over M1 in fibromyalgia (FM) patients has been examined by Cummiford et al. (Cummiford et al., 2016). Baseline connectivity between motor cortex and thalamic regions was used as a predictor of repetitive tDCS-generated analgesia (i.e., post-tDCS minus pre-tDCS clinical pain, visual analogue scale). In the seed to whole-brain analyses, greater pre-treatment connectivity between the left M1 seed and left ventral lateral (VL) thalamus, between the left primary somatosensory cortices seed and left anterior insula, and between the left VL thalamus seed and periaqueductal gray predicted greater improvement in clinical pain scores in patients with FM across sham and real tDCS periods.

Exploring fMRI during performance of a picture-naming task, Rosso and colleagues reported that functional connectivity from the right supplementary motor area to the right Broca’s area was significantly and positively correlated with tDCS-induced acceleration of picture naming (Rosso et al., 2014b), specifically effects of cathodal tDCS applied before task performance over Broca’s area.

Rosso and colleagues found another predictor explaining the variability of behavioral responses to cathodal tDCS, which was the inter-hemispheric functional balance between the right and left Broca’s area investigated via rs-fMRI connectivity maps (Rosso et al., 2014a). Four aphasic stroke patients with ischemic lesions involving the left Broca’s area were classified as responders, based on improvement in picture naming, after a single session of cathodal tDCS applied to the undamaged right Broca’s area in a double-blind randomized crossover trial, but seven participants were non-responders. Responders showed functional inter-hemispheric imbalance between the two Broca’s areas. Improvement in language performance after cathodal tDCS applied to the right undamaged Broca’s area in left middle cerebral artery stroke patients was observed when the left Broca’s area was damaged and there was a functional inter-hemispheric imbalance between the two Broca’s areas.

Unlike the previous functional MRI markers that predict only the behavioral response to tDCS, one study has used fMRI measures to predict neural responses measured by fMRI as well (Polanía et al., 2012). This study suggests that the motor cortex tDCS-induced functional connectivity alterations strongly depend on baseline intrinsic functional architecture of the human primary motor cortex (e.g., clustering coefficient and characteristic path-length).

##### 3.2.1.1. MPS for fMRI-as-Predictor

From the six experiments categorized in the “fMRI-as-predictor” scenario, four only used pre-tES fMRI, while the other two used pre- and post-tES fMRI. All studies used a crossover design. Four experiments were sham controlled trials and two had an active and sham control design. With regard to the number of tES sessions, five experiments applied stimulation in only one session, and one used a multiple-session study design (five consecutive days). Three different stimulation regions were used in “fMRI-as-predictor” experiments, which were the PFC (n=3), the sensorimotor cortex (n=2), and the parietal cortex (n=1). Stimulation intensity ranged from 1 to 2 mA, and intervention duration from 10 to 20 minutes. Three experiments used an rs-fMRI modality, one experiment used a task-based fMRI modality, one experiment used both DTI and rs-fMRI methods, and one experiment used both, DTI and task-related fMRI techniques. The most common predictors were functional connectivity markers (n=5 experiments), while cerebral activity markers were used only in one experiment (n=1).

#### 3.2.2. FMRI-as-Response

In the experiments categorized in the “fMRI-as-response” scenario, fMRI has been used to measure response to tES through predictive modeling (n=4 experiments). This scenario uses neuroimaging/behavioral predictors to explain neural outcomes in response to tES, as measured with fMRI. Lin and colleagues explored the potential utility of DTI to predict functional response (i.e., online minus pre-tDCS connectivity) to left DLPFC tDCS in 18 healthy subjects (Lin et al., 2017). Fractional anisotropy (FA), as the DTI metric of white matter tract integrity between the left DLPFC and left thalamus was computed using probabilistic tractography. Increased FA between the left DLPFC and left thalamus was associated with increased functional connectivity of these two regions after anodal tDCS. It also corresponded to regional cerebral blood flow (rCBF) changes in the activated voxels of the posterior insula in the sham condition during ongoing pain compared to the anodal condition, as assessed by ASL. Sankarasubramanian and colleagues investigated functional connectivity of the medial dorsal-DLPFC and ventroposterolateral-M1 before and after one session of active DLPFC tDCS to determine whether baseline pain thresholds could predict neural response (i.e., post-tDCS minus pre-tDCS connectivity) after a session of tDCS (Sankarasubramanian et al., 2017). They discovered that individuals with high baseline pain thresholds experience greater functional connectivity changes with active DLPFC tDCS, which highlights the role of the DLPFC in pain tolerance.

Polanía and colleagues examined the role of fMRI to measure the neural responses to tDCS in a regression modeling approach (Polanía et al., 2012). These investigators showed via rs-fMRI and graph theoretical functional connectivity analyses that the increase of the nodal clustering coefficient following left M1 cathodal tDCS strongly correlated with the baseline nodal clustering coefficient. They also reported a negative increase in the characteristic path length after left M1 anodal tDCS, which was positively correlated with baseline characteristic path length.

Abend and colleagues explored whether individual differences in neural response to medial PFC stimulation were associated with levels of depressive symptoms (Abend et al., 2018). They found that individuals with high baseline levels of depressive symptoms show increased stimulation-induced subgenual anterior-cingulate cortex activity while observing negative stimuli.

##### 3.2.2.1. MPS for fMRI-as-Response

The “fMRI-as-response” category explores the predictive power of baseline behavioral/neuroimaging markers in explaining the variance of neural outcomes in response to tDCS as measured with fMRI. The predictive behavioral/neuroimaging markers were pain thresholds (n=1 experiment), depressive symptom level (n=1 experiment), structural connectivity (mean FA; n=1 experiment), and functional connectivity (n=1 experiment; see Figure 7 for all predictive markers). All four studies used a crossover design. With regard to the control condition, two experiments had a sham controlled trial, and the other two experiments a mixed design (active and sham control designs). All “fMRI-as-response” experiments included experiments with single-session design only (n=4 experiments). Only two stimulation sites were probed, which were the PFC (n=3 experiments) and sensorimotor cortex (n=1 experiment). The duration of tES was 10 minutes and 20 minutes in the respective experiments. Three experiments used 1 mA intensity and one used 1.5 mA current intensity. Finally, with regard to the imaging modality, two experiments used rs-fMRI, one experiment used task-based fMRI, and one used a combination of ASL fMRI, and DTI.

### 3.3. Montage Experiments

In montage experiments, the location of the electrodes is defined based on functionally activated regions during a specific cognitive process. The aim is to look into the possible causal role of this area in the target cognitive process. Fourteen out of 220 experiments included in this systematic review are categorized as montage experiments. The main characteristics of these experiments are summarized in Table 1.

**Table 1.** Workflow and summary of montage studies employing tES to elucidate the causal role of certain brain regions involved in specific cognitive processes at the sites identified in the fMRI experiment.

| Montage Studies | Localization Phase |  |  |  | Targeting Phase |  |  |  |
| --- | --- | --- | --- | --- | --- | --- | --- | --- |
|  | Cognitive Function of Interest | Subjects | fMRI Task with Cognitive Function of Interest | Localization of Cognitive Function of Interest in Area X | Electrode Placement Targeting Area X | Subjects | tES Protocol Targeting Area X | Behavioral Task with Cognitive Function of Interest |
| (Ligneul et al., 2016) | Social dominance | n = 28 | Competitive task | rmPFC | Neuro-navigation system | n = 34 | rmPFC/Cz | Competitive task |
| (Zhang et al., 2016) | Intention-based cooperative decisions | n = 25 | Dictator game task | rLOFC | 10-20 EEG system | n = 42 | (Fp2/F2, F8, Fp1, lower eyelid)<br>(F2, F8, Fp1, lower eyelid/Fp2) | Dictator game task |
| (Mizuguchi et al., 2016) | Well-learned skillful motor performance | n = 15 | Finger-tapping task | Bilateral DLPFC, ACC, AIC, left cerebellar | 10-20 EEG system | n = 9* | F4/Fp1 | Finger-tapping task |
| (Hu et al., 2017) | Altruistic decision-making | n = 25 | Interactive dice game | mPFC, rDLPFC, rIPL | 10-20 EEG system | n = 114 | (F4/C4, FT8, Fp2, Fz)<br>(P4/C4, Pz, O2, P8) | Number comparison & Working memory task |
| (Wu et al., 2015) | Top-down and bottom-up attentional control | n = 35 | Majority function task | Bilateral rTPJ, rMOG, rFEF | 10-20 EEG system | n = 18 | Left cheek/(Between T4 and T6) | Majority function task |
| (Nihonsugi et al., 2015) | Intention-based economic decision | n = 41 | Trust game task | rDLPFC, ventral striatum, amygdala | 10-20 EEG system | n = 22 | F4/Oz | Trust game task |
| (Ashizuka et al., 2015) | Verbal politeness judgment | n = 20 | Sentence-judgment task | Precuneus | 10-20 EEG system | n = 12 | Fp2/Pz | Sentence-judgment task |
| (Woods et al., 2014) | Perceptual causality | n = 16 | Launching event task | Parietal, frontal | 10-20 EEG system | n = 16** | F3/F4<br>CP3/CP4 | Launching event task |
| (Xue et al., 2012) | Decision-making | n = 18 | Card-guessing task | Left LPFC | 10-20 EEG system, Neuro-navigation system | n = 18** | (Intersection of the F3–T3 line and the F7–C3 line) /Left cheek | Card-guessing task |
| (Clark et al., 2012) | Attentional process | n = 13 | Concealed object-learning task | rIFC, rPC | 10-20 EEG system | n = 83 | F10/Left upper arm<br>P4/Left upper arm | Concealed object-learning task |
| (Baker et al., 2010) | Language processing | n = 10 | Picture-naming task | Left frontal cortex | Neuro-navigation system | n = 10*** | Left frontal cortex/<br>Right shoulder | Picture-naming task |
| (Fischer et al., 2017) | Motor | n = 98 | M1 ROI analysis in rs-fMRI | Network of the left M1 hand area | 10-10 EEG system | n = 15 | C1, C2, C3, C4, T8/Fz, P3, P4 | NA |
| (Wang et al., 2017) | Updating the prediction in reinforcement learning | n = 81 | Iowa gambling task<br>Risk decision task | rACC/vmPFC, PCC | 10-10 EEG system | n = 68 | (Cz, Ex10, C2, FT10, Ex5, FC2/FCz, Fpz, Afz)<br>(Fz, TP7, O2, Pz, CPz/P8, FC6, FC5, O9) | Iowa gambling task<br>Risk decision task |
| (Frangou et al., 2018) | Perceptual decisions | n = 43 | Signal-in-noise task<br>Feature-differences task | Posterior occipito-temporal cortex | 10-20 EEG system | n = 84 | T6/Cz<br>Cz/T6 | Signal-in-noise task<br>Feature-differences task |

ACC: anterior cingulate cortex; AIC: anterior insular cortex; DLPFC: dorsolateral prefrontal cortex; LPFC: lateral prefrontal cortex; PCC: posterior cingulate cortex; mPFC: medial prefrontal cortex; rACC: rostral anterior cingulate cortex; rDLPFC: right dorsolateral prefrontal cortex; rFEF: right frontal eye field; rIFC: right inferior frontal cortex; rIPL: right inferior parietal lobe; rLOFC: right lateral orbitofrontal cortex; rMOG: right middle occipital gyrus; rmPFC: rostromedial prefrontal cortex; rPC: right parietal cortex; rTPJ: right temporoparietal junction; vmPFC: ventral medial prefrontal cortex.
\* Some of fMRI subjects participated in the tES experiment.
\*\* fMRI subjects were different from tES subjects.
\*\*\* All fMRI subjects participated in the tES experiment.

#### 3.3.1. MPS for Montage Experiments

There is significant heterogeneity in the MPS of montage experiments regarding imaging sequence type, fMRI timing, and paradigm design. With regard to fMRI characteristics, 13 experiments used a task-related fMRI design, and only one experiment used a resting-state design. All experiments relied on the BOLD signal for providing the spatial information needed to guide positioning of the tES electrodes.

In the fMRI-based tDCS-electrode localization stage, various tasks were administered to assess the cognitive function of interest, including competitive, dictator game, finger-tapping, interactive dice game, majority function, trust game, sentence-judgment, launching event, card-guessing, concealed object-learning, picture-naming, Iowa gambling, risk decision, signal-in-noise, and feature-differences tasks.

Extending the field from exploratory brain mapping (i.e., fMRI) to the causal inference aimed for by tES is demonstrated in Table 1. The most commonly targeted areas with tES based on the localization of the cognitive function of interest were the prefrontal (47.62%), followed by parietal (23.82%), sensorimotor (9.52%), temporal (9.52%), and occipital (9.52%) cortices.

For montage experiments, an important consideration is the methodology to optimize the reliability of the electrode placement for targeting of the desired brain regions, which included (1) 10-10 EEG system (14%, n=2), (2) 10-20 EEG system (64%, n=9), and (3) neuronavigation system (14%, n=2) approaches. One experiment used both the 10-20 EEG system and a neuronavigation system. Four experiments used multi-electrode tDCS intervention, while 10 experiments used square or rectangular electrodes for tES.

Thirteen experiments enrolled healthy subjects, and one experiment recruited stroke patients. Additionally, in 12 out of the 14 experiments, one participant group participated in the fMRI experiment, and another group in the tDCS experiment. One experiment employed fMRI and tES in the same group, and one experiment applied fMRI and tDCS on two separate groups, which had some overlap.

#### 3.3.2. Causal Interpretations derived from the fMRI-tES Combination

In a causal inference framework, neuroimaging-based mapping (i.e., fMRI) is performed before tES to derive the neural correlates associated with a behavioral state. tES is then applied to the respective area identified via mapping in the same group or in another group of subjects to interfere with the respective cognitive function of interest. Studies in this field include social dominance (Ligneul et al., 2016), motor (Mizuguchi et al., 2016), intention-based cooperative decision (Nihonsugi et al., 2015; Zhang et al., 2016), decision-making (Hu et al., 2017; Wang et al., 2017; Woods et al., 2014; Xue et al., 2012), attention (Clark et al., 2012; Wu et al., 2015), language (Ashizuka et al., 2015; Baker et al., 2010) and perceptual decision functions (Frangou et al., 2018).

One example study (Ashizuka et al., 2015) that used the methodology suggested in Table 1 tested the hypothesis that the precuneus is a fundamental hub involved in verbal politeness judgments. This study used fMRI to localize cortical areas that are involved in verbal politeness, which is a type of self-referential cognition that conforms to social hierarchy, and then used cathodal tDCS combined with the verbal politeness judgment task to characterize the crucial role of the precuneus in verbal politeness judgments. Another study (Woods et al., 2014) using similar procedures investigated the causal role of parietal versus frontal cortices involved in component processes of causal perception (i.e., spatial, temporal, and decision-making components). The authors used fMRI to identify neural substrates related to component processes important for perceptual causality and then tested the specific roles of the parietal and frontal cortices supporting perceptual causality with the use of tDCS that can probe the validity of these neural hypotheses. The results showed that the parietal cortex engages in causal perception via processing of spatial information, while the frontal cortex contributes to generalized decision-making processes. A third example study (Ligneul et al., 2016) demonstrated a causal role for the rostromedial prefrontal cortex in the emergence of social dominance based on competitive interactions, by combining computational modeling of behavior, identifying the pattern of neural activity with fMRI and tDCS guided by neuronavigation.

In most montage experiments, the reported effects were specific for a given task context and targeted brain area. Therefore, these experiments demonstrate that tES can relevantly help to establish causal evidence for a specific brain-behavior relationship, if researchers use a methodological procedure similar to that suggested in Table 1.

### 3.4. Replication within tES/fMRI Studies

Given the large parameter space of fMRI-tDCS studies, we asked whether similar studies could be conceivably combined to conduct a meta-analysis of a particular target region, taking into account both tES and fMRI parameters. As shown in Figure 8, experiments can be hierarchically organized, starting from a particular target area, and extending to paradigm, scanner and stimulation parameters. As one example, we focused on the sensorimotor cortex, which comprised the most frequent target among tDCS experiments overall, and sub-categorized the experiments by fMRI paradigm, electrode montage, and stimulation duration and intensity. At the final step, there are only five experiments which applied 1mA DC current for 10 minutes using a C3/Fp2 montage and looked at the effects using rs-fMRI (pre-post design). However, there are some minor but still important parameters which have not been reported in these five studies, such as the electrode orientation, and ramp up and ramp down duration at the start, and termination of the stimulation. This highlights the importance of careful reporting of all experimental details and lack of harmony and replications in the field.

**Figure 8:**
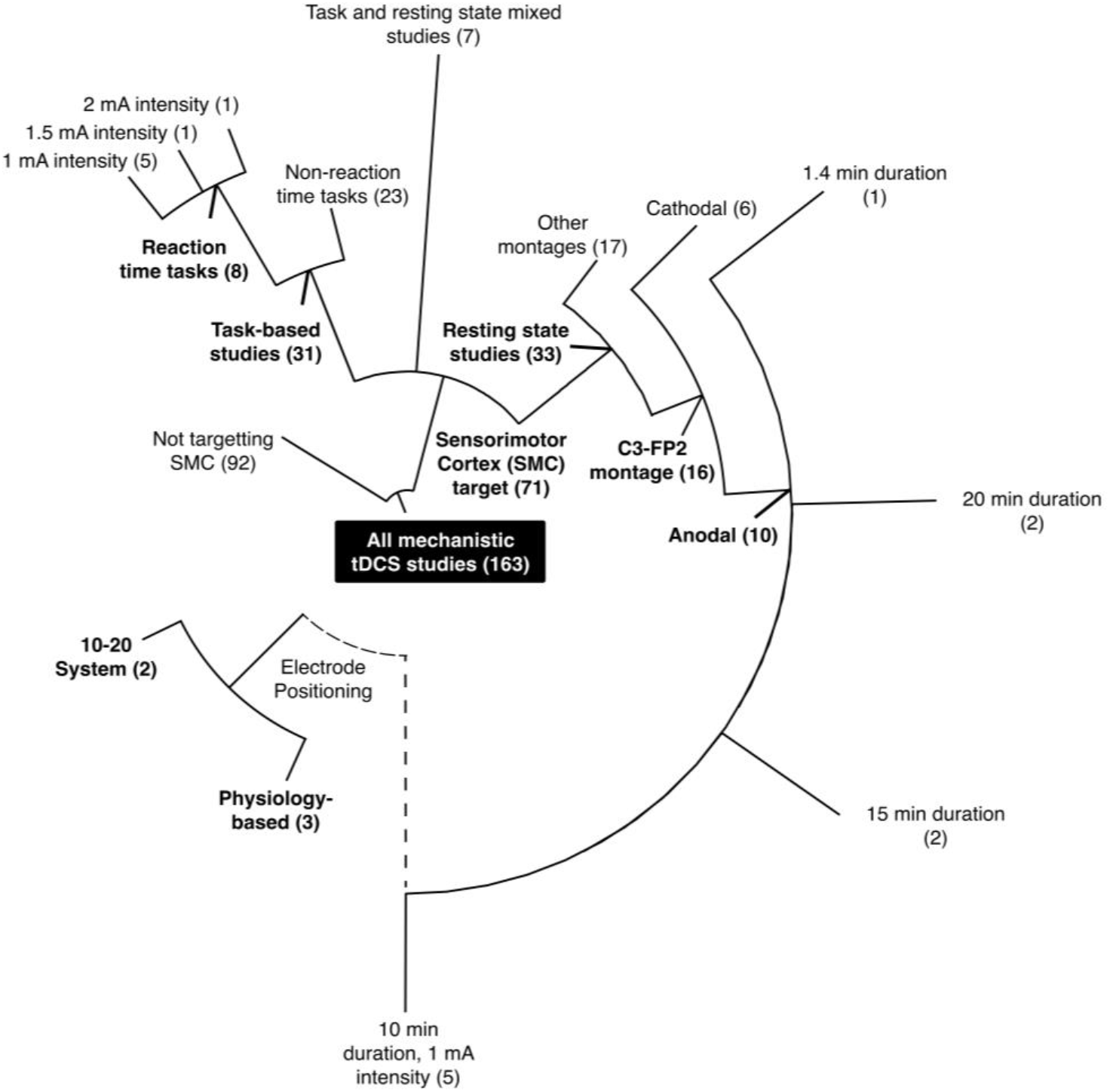
A sample hierarchical classification showing shrinkage in the number of experiments with matching experimental settings by adding experiment parameters. Starting from all 163 mechanistic tDCS-fMRI experiments, shown in the center, and categorizing them based on the stimulation target, type of fMRI, montage, and stimulation intensity and duration, we end up with a maximum of 3 experiments when grouping experiments by the most shared parameters (in this case, targeting the sensorimotor cortex (SMC) using a physiology-based C3/Fp2 montage with 1mA for 10 minutes, and recording resting state fMRI).

## 4. Discussion

In the present systematic review, we explored the methodological parameter space (MPS) of tES-fMRI studies published up to February 1^st^ 2019, and mapped the fMRI outcomes of respective studies across the brain. The MPS includes several important factors, such as electrode montage, tES intensity, duration, sham or control conditions, fMRI protocol, fMRI timing relative to tES, the specific behavioral task or clinical measure obtained. We have categorized these tES-fMRI studies into three main groups according to the role of fMRI: (1) mechanistic experiments; (2) predictive experiments; and (3) montage experiments. Out of 220 reviewed tES-fMRI experiments, 202 mechanistic experiments utilized fMRI as an objective measure to evaluate effects of stimulation on the targeted cortical region and remote areas. Ten experiments explored the power of fMRI maps as predictor/response for predicting neural/behavioral effects of tES. Fourteen tES-fMRI experiments utilized fMRI evidence to guide positioning of tES electrodes over brain networks/regions involved in a particular cognitive functions of interest. These 14 montage experiments explored the causal role of the targeted brain regions via modulating them with tES in the next step, and observing tES-driven changes of cognitive functions of interest. Overall, these findings provide a MPS framework for categorization of future fMRI-tES studies. We systematically showed a serious lack of overlap in the MPS between published tES-fMRI studies. This lack of overlap in the MPS between studies makes any meta-analysis and systematic/collective conclusion about mechanistic effects of tES complex. The findings from the reviewed studies highlighted several interesting trends, such as a large number of studies stimulating sensorimotor and prefrontal cortices for understanding mechanistic effects of tES, and reports of activation of extensive regions other than the target site by prefrontal stimulation. In the following sections, we will discuss the large methodological variability in tES-fMRI studies and provide recommendations to consider for future studies.

### 4.1. MPS-related Challenges and Gaps of Knowledge

Relevant sources of variability of tES effects might relate to the variability of methodological and design aspects employed in tES-fMRI studies. For example, with respect to methodological aspects, variations of tES protocols in terms of current intensity, duration, electrode size/shape and montage can alter stimulation effects. Beyond these parameters, an important source of variability of tES effects is the effect of incompletely explored nuances in methodological and technical considerations, such as the specific sham stimulation protocol, precision of electrode placements, subject handedness, and electrode conduction media, to name a few. In this section, we also provide some tentative guidelines that may help to control for the amount of variability, and thus will further strengthen experimental evidence, which will be relevant for future meta-analytic studies to elucidate the precise physiological and functional effects of tES.

#### 4.1.1. Variability of Sham Protocols

One important challenge in tES studies is to establish standardized methods to evaluate and increase the reliability/replicability of sham protocols, to ensure that stimulation effects can be ascribed specifically to tES. Most experiments included in this review (69%) used sham stimulation as the control condition, 21% had an active control, and 3% had a mixed design (both active and sham controls). In contrast, 7% of the experiments were performed without any control condition, and one study used a non-interventional design. Types of sham protocols were heterogeneous and included initial brief (+/-ramp) stimulation only (e.g., (D’Mello et al., 2017)), initial brief (+/-ramp) stimulation continued with low current stimulation with the same duration as real stimulation (e.g., (Miranda et al., 2009)), and initial and final brief (+/-ramp) stimulation (e.g., (Palm et al., 2013)). Efforts to improve the quality of sham reliability are ongoing (Fonteneau et al., 2019; Garnett and Den Ouden, 2015; Kessler et al., 2012). Future tES-fMRI studies should utilize a standardized reliable sham protocol. In this consideration, it would be important to rule out whether respective sham protocols induce stimulation-dependent physiological effects, as is likely the case with extended low-intensity stimulation. These factors will contribute to poor blinding since such low intensities of tACS will not induce somatosensory or other perceptions but may still induce physiological effects (Loo et al., 2018).

#### 4.1.2. Reproducibility of Electrode Placements

Reproducibility of electrode placements is another not completely solved frontier. Even slight differences of electrode positions can lead to significant differences in where and how much current is delivered to the respective brain regions (Kessler et al., 2013; Minhas et al., 2012; Nitsche and Paulus, 2000; Woods et al., 2015). For example, Nitsche and Paulus showed that differences in electrode placement critically determined whether tDCS affected TMS-generated MEPs (Nitsche and Paulus, 2000). Computational modeling can be a powerful tool to assess differences between relative positions of tES electrodes with respect to the targeted brain area. In one example, Woods and colleagues developed an MRI-derived finite element model to demonstrate that as little as a 5% drift in electrode locations can significantly change the predicted electric field in the brain as well as the intensity of stimulation in specific brain regions (Woods et al., 2015). As head size, shape and anatomy varies greatly across individuals (Woods et al., 2016), it is crucial to select the appropriate strategy to localize the electrode placement. The localization of electrode position in those experiments which reported the respective approach was determined either using the 10-20 (or 10-10) electrode placement system/distance-based system (72%), physiology-based placement (15%), neuro-navigation systems (5%) or the 10-20 EEG system combined with anatomical landmarks/neuro-navigation systems (3%). It should be pointed out that physiology-based electrode placement can only be accomplished for motor and primary sensory cortices; however, further developments in this direction are still required, and might change this situation (e.g., TMS-EEG methods). Future tES-fMRI studies should strive to use physiological methods, or neuro-navigation based methods instead of purely scalp-measurement based systems (e.g., the 10-20 EEG system), to ensure reliable electrode positioning and potential for future replication.

#### 4.1.3. Right vs Left Handedness

Only one of the included studies (Kasahara et al., 2013), took the importance of handedness into account for the interpretation of the results, despite the consideration that non-fMRI tES and TMS studies have identified handedness as a potential confound (Kasuga et al., 2015; Van den Eynde et al., 2012). Hemispheric dominance might be a major source of variability, as around 95–99 percent of right-handed individuals are left-hemisphere dominant for language, whereas about 70 percent of left-handed individuals are left-hemisphere dominant for language (Corballis, 2014). In addition, handedness might be of clinical significance, since left-handed people are more at risk for addictive disorders (Sperling et al., 2000), breast cancer (Fritschi et al., 2007), allergies and autoimmune disorders (Geschwind and Behan, 1982), autism (Colby and Parkison, 1977), and other diseases. Assessing the hemispheric dominance by means of a special questionnaire focused on handedness laterality (Oldfield, 1971) in subjects included in brain stimulation studies might provide statistically useful information about brain functioning.

#### 4.1.4. Contact Medium: Gel vs. Saline vs. Conductive Paste

Another critical consideration for tES is optimizing the contact medium of the metal or conductive rubber electrode with the skin. The electrolyte used for conduction can be applied in a sponge pocket covering the electrode (i.e., saline) (DaSilva et al., 2011) or, in the case of conductive gel, applied directly on the skin. For saline, oversaturation of the sponge pouches remarkably undermines the ability to perform reproducible tES application and obtain reliable results. If these sponges are over-soaked, saline may have a tendency to spread beyond the electrode region at the level of the scalp, causing changes in the spread and direction of current flow, and undermining tES replicablility (Woods et al., 2016). Use of methods to quantify saline (e.g., syringes) can ensure a consistent and appropriate amount of contact medium. Alternatively, electro-conductive gel under the surface of the electrode/sponge also improves stability of the scalp contact, providing uniform current distribution, especially for participants with dense hair (Woods et al., 2016). However, if gel is applied to the base of the rubber electrode, the evaporation of gel may occur as a result of increased temperatures as well as prolonged stimulus delivery times, which increases the risk of burns to the scalp (Lagopoulos and Degabriele, 2008). Nevertheless, not all gel types are equivalent, and may induce different cutaneous sensations in volunteers, especially viscous gels, that are also difficult to apply to the base of the rubber electrode (Fertonani et al., 2015; Poreisz et al., 2007; Saturnino et al., 2015). Similarly, for prolonged MRI sessions, conductive pastes are appropriate as a contact medium for concurrent tES-fMRI, as they avoid premature drying of electrode sponges when using saline. Care must also be taken as excessive paste may smear, increasing the contact area beyond the electrode placement, changing expected current density, and potentially creating MRI-hazardous short circuits (Esmaeilpour et al., 2019). Drying of the sponges results in a decrease of the conductance of the electrode-skin interface, which may increase the risk of pain and skin burning to the subject. If conductive pastes are used, the thickness of the paste should be consistent (~3 mm) and should cover the electrode completely in order to ensure that stimulation is delivered evenly across the electrode (Woods et al., 2016). Among those experiments which reported the contact medium (90%), 53% of experiments used a saline-based contact medium, 25% used a conductive paste, and 12% used an EEG gel. Finally, the type of contact medium should be selected according to the methodology of the study (i.e, offline or online tES-fMRI), as well as the inherent design constraints (e.g., oversaturation of sponges, evaporation of saline/drying of electrode gel, etc.).

### 4.2. Towards Harmonization and Replication in MPS

Despite these challenges, there are specific suggestions to resolve these issues for future tES-fMRI studies. These suggestions include standardizing sham and active control interventions, optimizing stimulation and imaging parameters, and standardizing clinical measures. Such solutions can help to optimize stimulation protocols and further strengthen comparability between studies and set the foundation for future meta-analytic approaches to elucidate the efficacy, reliability, and mechanistic foundations of tES. For this, it is important to minimize methodological heterogeneity in order to achieve a better understanding of tES as a promising basic research and therapeutic approach. Finally, given the large and increasing number of research laboratories using tES-fMRI, we recommend development of a tES/fMRI consortium committed to aggregating and sharing databases and experimental protocols across research groups.

One of the major missions of this international network/consortium will be to increase replicability of published evidence in the field. Concerns have been raised about the reproducibility of scientific publications with reports that the results of experiments in many domains of science could not be replicated (Begley, 2013; Begley and Ioannidis, 2015; Science, 2015). When designing a study, it is important to provide sufficiently detailed methodological and analytical information so that methods can be precisely repeated. Additionally, of similar importance, there should be agreement about the level and nature of processing of the data, the level of detail in the description of measurement methods, and the completeness of the final analytical results. It is important to note that such an agreement does not exist for tES-fMRI studies. To clarify this issue, a standard checklist including all decisive steps and parameters should be developed for reporting and interpreting of tES-fMRI studies. Such a checklist will improve evaluation of the methodological reproducibility of results and eventual development of useful methodological standardization with real-world applicability.

### 4.3. tES/fMRI Mechanistic Experiments

Despite the fact that numerous studies have used tES combined with fMRI to resolve mechanistic questions, the large space of stimulation and imaging parameters, which differs considerably across studies, does not allow one to use meta-analytic procedures to combine all of these studies and derive an estimate of the effect size. Such analyses can only be applied within coherent sets of studies which have used the same parameters (Nitsche et al., 2015). For the studies included in the present review, it was not possible to identify more than five studies using the same experimental design, task paradigm, and stimulation parameters (Figure 8). Thus, a major challenge in the field is to address the reliability of experimental results, given that 1) variations of stimulation intensity, duration and montage can impact the stimulation effects, even in a non-linear way (Batsikadze et al., 2013; Jamil et al., 2016; Monte-Silva et al., 2013), and 2) studies involving NIBS have shown high levels of inter-individual variability, which has so far not been explored systematically with respect to its sources (Ridding and Ziemann, 2010). Methodological aspects are also relevant with respect to reproducibility. Beyond the factors mentioned above, online tES-fMRI studies are more prone to artefactual noise than other scenarios, which stresses the importance of having properly placed and shielded electrode cables and stimulation equipment within the scanning area (Antal et al., 2014; Woods et al., 2016). Additionally, subject-specific aspects play a crucial role, such as differences in arousal, anxiety, or attentional state, which can fluctuate considerably when subjects enter the fMRI scanner (van Minde et al., 2013) or when asked to remain awake for a prolonged period of time while lying in a supine position (Tagliazucchi and Laufs, 2014). Finally, the link between task-based behavioral findings and physiological measures, such as changes in connectivity or perfusion, may not always be straightforward. For example, improved performance during motor learning tasks is known to be associated not only with immediate activity enhancements of task-relevant areas, but also with delayed activity reductions of the respective motor cortical networks (Lin et al., 2012; Pascual-Leone et al., 1994). Such findings do not invalidate the functional role of these networks, rather they may reflect an increase in the selectivity of task-relevant networks (Moisa et al., 2016). TDCS, and even other NIBS protocols may affect physiological and behavioral read-outs in diverse ways during different stages of learning, for example in visuomotor coordination (Antal et al., 2004; Polanía et al., 2018), which has to be taken into account when interpreting results.

In order to address and overcome these challenges, we propose some tentative guidelines on the planning and execution of tES-fMRI studies, which may help to reduce the amount of experimental variability and further strengthen reliability and validity of results in order for future meta-analytic studies to be able to elucidate the precise physiological and functional effects of tES.

1. One major strength brought about by combining tES and neuroimaging techniques is the potential to look at the link between physiological and behavioral data. However, in our review of the extant studies, we observed that the majority of studies which reported both behavioral and physiological findings did not report the correlation between these findings. This point needs to be considered in future studies.
2. More attention should be devoted to the precise reporting of methods, protocols and results to allow more accurate interpretation and future summary of the data. This includes reporting of the electrode shape, size, orientation, location on the scalp, current intensity, stimulation duration, fade-in/fade-out times and behavioral state of subjects during stimulation (Bikson et al., 2019). A standard checklist including all of the decisive steps and parameters can help to make future experimental settings more homogeneous.
3. The placement of electrodes over target sites should be precisely done, and we recommend the use of physiological methods or neuro-navigation based methods over purely scalp-measurement based systems (e.g., the 10-20 EEG system).
4. For task-based studies, a control condition should be included in order to be able to assess whether the behavioral effects as a result of tES are specific to the task.
5. Careful avoidance of stimulation-related artifacts for online tES-fMRI studies is of paramount importance.
6. Using a pre-post intervention design when available, conditional on not moving the subjects out of the scanner for the respective assessments, in order to reduce additional variability and non-specific effects resulting from body movement into and out of the scanner is advantageous.
7. Considering the critical non-linear and therefore not-easily predictable role of different stimulation parameters, titration studies are required to systematically investigate the effect of each parameter on different levels of brain function, including neurochemical, neuroelectrical, and oscillation changes.
8. The potential of multimodal neuroimaging approaches for understanding human cognition has not been fully exploited, yet. Combined EEG-fMRI studies provide data with high resolution in both temporal and spatial dimensions which can be helpful to achieve a better understanding of the underlying mechanisms (Abreu et al., 2018).
9. Computational models can be used to design individually optimized stimulation configurations to target a special brain area. On the other hand, data obtained from tES-fMRI studies could be used to estimate the predictive power of computational models to explain the inter and intra individual variation in fMRI response to tES.
10. Concurrent acquisition of fMRI with tES has to be done in a methodologically rigorous way, as field artifacts might alter MRS and ASL-based sequences, and even lead to recording fake BOLD signals (false positive functional activities) (Antal et al., 2014; Esmaeilpour et al., 2019). It is advised to conduct a standard procedure to assure image quality. One suggested method to prevent recording false data is acquisition of EPI field maps for a short duration of time under two conditions, with and without stimulation, and then to compare these (Woods et al., 2016).

### 4.4. tES/fMRI Predictive Experiments

This systematic review suggests that functional and structural neuroimaging approaches might be able to predict the likelihood of behavioral and/or neural responses to tES interventions, and eventually provide innovative strategies to advance the field towards precision medicine. Taken together, these findings provide preliminary but supporting evidence that fMRI might help to disentangle the variability of tES to effects into inter- and intra-individual sources. tES-response prediction can be a crucial element of validating fMRI biomarkers. Some predictive experiments indicate that alteration of brain functional connectivity measured with fMRI is associated with a large variety of neurological and psychiatric disorders and therefore promises to be a potential biomarker for predicting treatment response to tDCS (Cavaliere et al., 2016; Cummiford et al., 2016). In order to fulfill this promise, it is important to address a range of challenges, including methodological, interpretation, and translational aspects. Surprisingly, there are no established biomarkers available for predicting tES response in a large variety of brain disorders. Efforts in this direction should be intensified. One potential marker with a potential clinical value for tDCS response is disease-related abnormal functional connectivity in corticothalamic and extrinsic control networks (Cavaliere et al., 2016; Cummiford et al., 2016). These fMRI markers have been considered as predictors of tDCS response in the assessment of consciousness in MCS patients and have also shown promise for reducing pain in fibromyalgia patients.

#### 4.4.1. Challenges and Perspectives

Considerable methodological challenges need to be addressed with respect to predictive tES-fMRI studies. Most studies apply correlation or regression models for establishing brain–behavior relationships using neuroimaging data. As mentioned in the results section, these methods often tend to overfit the data and, as a result, limit generalizability. Most of the predictive experiments did not use a cross-validation approach, which makes it difficult to assess the generalizability of the results. Kriegskorte et al. (Kriegeskorte et al., 2009) report that circularity in selection and selective analyses yield spuriously significant test results. The use of appropriate cross-validation approaches to ensure statistical independence between feature selection and regression/classification is warranted in future studies, thus eliminating distorted descriptive statistics and invalid statistical inferences.

While predictive biomarkers are encouraging on the whole, a key aspect in considering their clinical potential is whether these can be employed for prediction at the individual level to a clinically significant degree of accuracy. In clinical decisions, predictive biomarkers require analytical validity, clinical validity, and clinical utility. Although the achievement of analytical and clinical validity is relatively free from ambiguity, attaining clinical utility is highly challenging. It is well-established that predictive biomarkers have clinical applicability if they are actionable for making treatment decisions in a way that leads to the patient’s benefit (Group, 2016). Predictive biomarkers provide information on the likelihood of response to treatment (Walther et al., 2009) and support the procedure of clinical decisions (Voon and Kong, 2011). Pilot studies in the field of clinical tES application deliver initial promising results. It was shown that the difference between responsive and non-responsive MCS patients to tDCS over the left DLPFC is related to resting state DLPFC-IFG functional connectivity (Cavaliere et al., 2016). Furthermore, differences in the anatomical integrity of the arcuate fasciculus, as assessed by the FA ratio, is associated with tDCS treatment response in aphasic stroke patients (Rosso et al., 2014a). These studies show that proper classification of responders and non-responders might be possible on the basis of structural and functional neuroimaging measures. One promising direction is the application of machine learning techniques for neuroimaging measures, which might have potential for individual-level prediction of tES response for various disorders. Previous studies confirm its potential for predicting responses to diverse intervention approaches, including cognitive behavioral therapy (Costafreda et al., 2009b; Fu et al., 2008), TMS (Avissar et al., 2017; Drysdale et al., 2017), and medication (Costafreda et al., 2009a).

An open question for tES-response prediction is whether the structural morphometric or connectivity variances underlie the functional and behavioral responses to tES. It is likely that the structural integrity of white matter tracts is predictive not only of the behavioral but also of the physiological response to tES. Using a baseline DTI measure, Lin and colleagues reported an association between the individual left DLPFC-thalamic tract integrity, as measured by the mean FA of the tract, and anodal tDCS-induced changes of functional connectivity between the left DLPFC and left thalamus (Lin et al., 2017). Specifically, the individuals with the highest structural integrity of the left DLPFC-thalamic tract showed the highest functional coupling between these two regions during anodal tDCS. Overall, this suggests the dependence of physiological and behavioral effects of tES on the structural measures of the relevant interconnections. Thus, pre-stimulation structural neuroimaging measures might be valuable to elucidate reasons for variations in the efficacy of tDCS.

To explore how maximal tES efficacy is affected by inter-individual variability, which restricts whole group effect size (Ammann et al., 2017; Wiethoff et al., 2014), and to improve dose optimization strategies (Kessler et al., 2013), future studies should combine current flow modeling with fMRI measures. Computational models of current flow can be used to retrospectively address the role of brain current flow according to the individual anatomy and tissue architecture in tES response variability. For such a predictive framework, a non-trivial question is to what degree variability in electric field distribution/intensity based on computational finite element method (FEM) models can predict variability of the neuronal, and ultimatively behavioral/psychological response to tES. However, there is no fMRI study available which has used computational FEM models alone for predicting neural responses of tES (Figure 7; hypothetical study). This might, however, be a critical issue for further research to understand the impact of tES on the neuronal level.

### 4.5. tES/fMRI Montage Experiments

The goal of using fMRI for determining electrode montage is to study causal brain-behavior relationships in humans with fMRI-informed tES. In these studies, fMRI is used in order to determine the desired cortical region underlying specific cognitive processes. Overall, results from 14 montage experiments in our systematic review show that tES can modulate different aspects of cognitive processing targeting brain areas in frontal, parietal, temporal, sensorimotor, and occipital cortices.

The overarching hypothesis of these studies is that tES applied to the candidate brain regions identified by fMRI will affect defined cognitive processes (Baker, Rorden and Fridriksson, 2010; Xue *et al*., 2012; Clark *et al*., 2012; Woods *et al*., 2014; Ashizuka *et al*., 2015; Wu *et al*., 2015; Nihonsugi, Ihara and Haruno, 2015; Zhang *et al*., 2016; Ligneul *et al*., 2016; Mizuguchi *et al*., 2016; Wang *et al*., 2017; Fischer *et al*., 2017; Hu *et al*., 2017; Frangou *et al*., 2018). Findings from the reviewed studies suggest indeed that the respective frontal, parietal, temporal, sensorimotor, and occipital cortices have causal relevance for specific cognitive processes, as shown in Table 1.

#### 4.5.1. Challenges and Perspectives

There are important challenges to be addressed in the “causal inference” approach using fMRI-informed tES. These challenges relate to variability of methodological procedures (localization of electrode position, sample size, control task, etc.) employed in montage experiments. This variability indicates a lack of explicit guidelines in how to design tES-fMRI studies to enable them to deliver conclusive evidence for a specific, causally defined brain behavior relationship. This includes details of fMRI employment to guide tES electrode sites, and employment of tES to provide causal inferences.

Extending the field from exploratory brain mapping (i.e., fMRI) to brain stimulation approaches (i.e., tES) requires careful consideration of all steps of a montage study design (Table 1). When designing the respective experiments, it is necessary to precisely define the brain region of interest to receive tES, and the cognitive process that should be targeted. This last step requires careful planning to design a cognitive task (both during and after fMRI) that taps into the cognitive function of interest to establish the brain region or network that should be stimulated. As the electrode placement to target the desired brain region varies across individuals (due to differences in head size and shape (Woods et al., 2016)), it is important to use a method to optimize the reliability of the localization of the respective electrode positions. A methodological approach that contributes to this limitation is that most montage studies (approximately 57%) used the 10-20 EEG system for localization of electrode position and thus, there might be variability across subjects. The use of neuro-navigation in order to more precisely target the tES region of interest in each subject, based on individual functional/anatomical MRI evidence, paves the way to derive the coordinates for which brain networks are maximally involved in a particular cognitive task, and is likely suited to compensate for relevant sources of interindividual variability. To this aim, either (1) the coordinates are recognized in prior group studies and then denormalized to appraise the position in the particular individual’s brain, for example for the rmPFC (Ligneul et al., 2016), or (2) independent fMRI experiments are performed to estimate the coordinates associated with each subject (Baker et al., 2010). Given the large interindividual anatomical variability, the latter approach will likely provide maximal precision and is feasible for all potential target regions. Additionally, simultaneous application of NIBS during concurrent neuroimaging allows dynamic monitoring of task-related brain networks that operate either during or after an intervention, thereby allowing study of causal links between brain areas/ networks and specific aspects of behavior. This makes it possible to spatially adjust the stimulation electrodes to the relevant brain site. However, it is important to note that the quick relocation of stimulation electrodes is not possible in tES studies. However, MR-compatible multi-channel or switch-matrix stimulators are offered in the future to target specific brain networks with multi-focal targets. Additionally, it might become feasible to automatically configure optimal montages based on the current brain state in future studies. It should be noted that a transformation function that can reasonably predict fMRI signal change due to a certain tES stimulation protocol in the individual level is not yet developed.

Furthermore, to ensure robust interpretation of the data and increase the potential for future replication, tES-fMRI studies should strive to adhere to sample sizes appropriate for tES and fMRI studies (average sample size of all tES-fMRI studies published between 2000 and February 1st, 2019 = 24.2 ± 20.9), based on power calculations. Studies with sample sizes of n<20 are probably at risk of being irreproducible (Simonsohn et al., 2014).

### 4.6. Conclusion

We have provided an overview of the methodological aspects of tES integration with functional MR imaging in order to develop a categorization of the methodological parameter space of these studies. tES protocols may be enhanced by functional MR imaging. First, fMRI can provide proxy measures for the potential neuronal mechanisms underlying tES effects after (offline) or during (online) stimulation (i.e., mechanistic experiments). Secondly, fMRI can be used in predicting neural/behavioral response to tES (i.e., predictive experiments). Thirdly, fMRI can guide tES with regard to where to apply the stimulation (i.e., montage experiments). While hypotheses about the neural correlates related to specific cognitive processes can be generated before tES, those hypotheses need to be tested to enable the causal contribution of candidate brain regions to a specific cognitive or behavioural process. Future research plans should further develop the methodological parameter space to improve the precision and efficiency of tES-fMRI applications for neuroscience research systems and to exploit the therapeutic benefits of combining tES with functional neuroimaging. Developing a transformation function to reasonably predict fMRI response to a certain tES montage and a protocol in both temporal and spatial dimensions at the individual level based on the accumulating harmonized evidence and machine learning methods is an exciting goal for the future of tES-fMRI studies in the next decades.

## Data Availability

https://osf.io/5jbtg/?view_only=027124979b144c96bf220a98a6896e39

## Data and code availability statements

The tES-fMRI database is available at

(https://osf.io/5jbtg/?view_only=027124979b144c96bf220a98a6896e39).

## CRediT authorship contribution statement

**Peyman Ghobadi-Azbari:** Conceptualization, Methodology, Formal analysis, Investigation, Writing -Original Draft, Writing - Review & Editing, Visualization. **Asif Jamil:** Conceptualization, Formal analysis, Investigation, Writing - Original Draft, Writing - Review & Editing, Visualization. **Fatemeh Yavari:** Conceptualization, Formal analysis, Writing - Original Draft, Writing - Review & Editing, Visualization. **Zeinab Esmaeilpour:** Conceptualization, Writing - Review & Editing. **Nastaran Malmir:** Investigation. **Rasoul Mahdavifar-Khayati:** Writing - Review & Editing. **Ghazaleh Soleimani:** Writing - Review & Editing. **Yoon-Hee Cha:** Writing - Review & Editing. **A. Duke Shereen:** Writing - Review & Editing. **Michael A. Nitsche:** Conceptualization, Writing - Review & Editing. **Marom Bikson:** Conceptualization, Writing - Review & Editing. **Hamed Ekhtiari:** Conceptualization, Methodology, Writing - Original Draft, Writing - Review & Editing, Visualization, Supervision.

## References

Abellaneda-Pérez, K., Vaqué-Alcázar, L., Perellón-Alfonso, R., Bargalló, N., Kuo, M.-F., Pascual-Leone, A.Nitsche, M.A., Bartrés-Faz, D., 2019. Differential tDCS and tACS Effects on Working Memory-Related Neural Activity and Resting-State Connectivity. Front. Neurosci. 13, 1440. https://doi.org/10.3389/fnins.2019.01440

Abend, R., Sar-El, R., Gonen, T., Jalon, I., Vaisvaser, S., Bar-Haim, Y., Hendler, T., 2018. Modulating Emotional Experience Using Electrical Stimulation of the Medial-Prefrontal Cortex: A Preliminary tDCS-fMRI Study. Neuromodulation. https://doi.org/10.1111/ner.12787

Abreu, R., Leal, A., Figueiredo, P., 2018. EEG-informed fMRI: A review of data analysis methods. Front. Hum. Neurosci. https://doi.org/10.3389/fnhum.2018.00029

Ammann, C., Lindquist, M.A., Celnik, P.A. %J B. stimulation, 2017. Response variability of different anodal transcranial direct current stimulation intensities across multiple sessions 10, 757–763.

Antal, A, Alekseichuk, I., Bikson, M., Brockmoller, J., Brunoni, A.R., Chen, R., Cohen, L.G., Dowthwaite, G., Ellrich, J., Floel, A., Fregni, F., George, M.S., Hamilton, R., Haueisen, J., Herrmann, C.S., Hummel, F.C., Lefaucheur, J.P., Liebetanz, D., Loo, C.K., McCaig, C.D., Miniussi, C., Miranda, P.C., Moliadze, V., Nitsche, M.A., Nowak, R., Padberg, F., Pascual-Leone, A., Poppendieck, W., Priori, A., Rossi, S., Rossini, P.M., Rothwell, J., Rueger, M.A., Ruffini, G., Schellhorn, K., Siebner, H.R., Ugawa, Y., Wexler, A., Ziemann, U., Hallett, M., Paulus, W., 2017. Low intensity transcranial electric stimulation: Safety, ethical, legal regulatory and application guidelines. Clin. Neurophysiol. 128, 1774–1809. https://doi.org/10.1016/j.clinph.2017.06.001

Antal, A., Alekseichuk, I., Bikson, M., Brockmöller, J., Brunoni, A.R., Chen, R., Cohen, L.G., Dowthwaite, G., Ellrich, J., Flöel, A., Fregni, F., George, M.S., Hamilton, R., Haueisen, J., Herrmann, C.S., Hummel, F.C., Lefaucheur, J.P., Liebetanz, D., Loo, C.K., McCaig, C.D., Miniussi, C., Miranda, P.C., Moliadze, V., Nitsche, M.A., Nowak, R., Padberg, F., Pascual-Leone, A., Poppendieck, W., Priori, A., Rossi, S., Rossini, P.M., Rothwell, J., Rueger, M.A., Ruffini, G., Schellhorn, K., Siebner, H.R., Ugawa, Y., Wexler, A., Ziemann, U., Hallett, M., Paulus, W., 2017. Low intensity transcranial electric stimulation: Safety, ethical, legal regulatory and application guidelines. Clin. Neurophysiol. 128, 1774–1809. https://doi.org/10.1016/j.clinph.2017.06.001

Antal, A., Bikson, M., Datta, A., Lafon, B., Dechent, P., Parra, L.C., Paulus, W., 2014. Imaging artifacts induced by electrical stimulation during conventional fMRI of the brain. Neuroimage 85, 1040–1047. https://doi.org/10.1016/j.neuroimage.2012.10.026

Antal, A., Nitsche, M. a, Kruse, W., Kincses, T.Z., Hoffmann, K.-P., Paulus, W., 2004. Direct current stimulation over V5 enhances visuomotor coordination by improving motion perception in humans. J. Cogn. Neurosci. 16, 521–527. https://doi.org/10.1162/089892904323057263

Antonenko, D., Schubert, F., Bohm, F., Ittermann, B., Aydin, S., Hayek, D., Grittner, U., Flöel, A., 2017. tDCS-Induced Modulation of GABA Levels and Resting-State Functional Connectivity in Older Adults. J. Neurosci. 37, 4065–4073.

Ashizuka, A., Mima, T., Sawamoto, N., Aso, T., Oishi, N., Sugihara, G., Kawada, R., Takahashi, H., Murai, T., Fukuyama, H., 2015. Functional relevance of the precuneus in verbal politeness. Neurosci. Res. 91, 48–56.

Avissar, M., Powell, F., Ilieva, I., Respino, M., Gunning, F.M., Liston, C., Dubin, M.J., 2017. Functional connectivity of the left DLPFC to striatum predicts treatment response of depression to TMS. Brain Stimul. 10, 919–925. https://doi.org/10.1016/j.brs.2017.07.002

Bachinger, M., Zerbi, V., Moisa, M., Polanía, R., Liu, Q., Mantini, D., Ruff, C., Wenderoth, N., 2017. Concurrent tACS-fMRI Reveals Causal Influence of Power Synchronized Neural Activity on Resting State fMRI Connectivity. J. Neurosci. 37, 4766–4777. https://doi.org/10.1523/JNEUROSCI.1756-16.2017

Baker, J.M., Rorden, C., Fridriksson, J., 2010. Using transcranial direct-current stimulation to treat stroke patients with aphasia. Stroke 41, 1229–1236.

Barbati, S.A., Cocco, S., Longo, V., Spinelli, M., Gironi, K., Mattera, A., Paciello, F., Colussi, C., Podda, M.V., Grassi, C., 2019. Enhancing Plasticity Mechanisms in the Mouse Motor Cortex by Anodal Transcranial Direct-Current Stimulation: The Contribution of Nitric Oxide Signaling. Cereb. Cortex. https://doi.org/10.1093/cercor/bhz288

Barker, A.T., Jalinous, R., Freeston, I.L., 1985. Non-invasive magnetic stimulation of human motor cortex. Lancet (London, England) 1, 1106–7.

Batsikadze, G., Moliadze, V., Paulus, W., Kuo, M.-F., Nitsche, M.A., 2013. Partially non-linear stimulation intensity-dependent effects of direct current stimulation on motor cortex excitability in humans. J. Physiol. 591, 1987–2000. https://doi.org/10.1113/jphysiol.2012.249730

Bikson, M., Brunoni, A.R., Charvet, L.E., Clark, V.P., Cohen, L.G., Deng, Z. De, Dmochowski, J., Edwards, D.J., Frohlich, F., Kappenman, E.S., Lim, K.O., Loo, C., Mantovani, A., McMullen, D.P., Parra, L.C., Pearson, M., Richardson, J.D., Rumsey, J.M., Sehatpour, P., Sommers, D., Unal, G., Wassermann, E.M., Woods, A.J., Lisanby, S.H., 2018. Rigor and reproducibility in research with transcranial electrical stimulation: An NIMH-sponsored workshop. Brain Stimul. https://doi.org/10.1016/j.brs.2017.12.008

Bikson, M., Esmaeilpour, Z., Adair, D., Kronberg, G., Tyler, W.J., Antal, A., Datta, A., Sabel, B.A., Nitsche, M.A., Loo, C., Edwards, D., Ekhtiari, H., Knotkova, H., Woods, A.J., Hampstead, B.M., Badran, B.W., Peterchev, A. V., 2019. Transcranial electrical stimulation nomenclature. Brain Stimul. https://doi.org/10.1016/j.brs.2019.07.010

Bikson, M., Grossman, P., Thomas, C., Zannou, A.L., Jiang, J., Adnan, T., Mourdoukoutas, A.P., Kronberg, G., Truong, D., Boggio, P., Brunoni, A.R., Charvet, L., Fregni, F., Fritsch, B., Gillick, B., Hamilton, R.H., Hampstead, B.M., Jankord, R., Kirton, A., Knotkova, H., Liebetanz, D., Liu, A., Loo, C., Nitsche, M.A., Reis, J., Richardson, J.D., Rotenberg, A., Turkeltaub, P.E., Woods, A.J., 2016. Safety of Transcranial Direct Current Stimulation: Evidence Based Update 2016. Brain Stimul. 9, 641–661. https://doi.org/10.1016/j.brs.2016.06.004

Bikson, M., Name, A., Rahman, A., 2013. Origins of specificity during tDCS: anatomical, activity-selective, and input-bias mechanisms. Front. Hum. Neurosci. 7, 688. https://doi.org/10.3389/fnhum.2013.00688

Brunoni, A.R., Sampaio-Junior, B., Moffa, A.H., Aparício, L. V., Gordon, P., Klein, I., Rios, R.M., Razza, L.B., Loo, C., Padberg, F., Valiengo, L., 2019. Noninvasive brain stimulation in psychiatric disorders: a primer. Brazilian J. Psychiatry 41, 70–81. https://doi.org/10.1590/1516-4446-2017-0018

Buch, E.R., Santarnecchi, E., Antal, A., Born, J., Celnik, P.A., Classen, J., Gerloff, C., Hallett, M., Hummel, F.C., Nitsche, M.A., Pascual-Leone, A., Paulus, W.J., Reis, J., Robertson, E.M., Rothwell, J.C., Sandrini, M., Schambra, H.M., Wassermann, E.M., Ziemann, U., Cohen, L.G., 2017. Effects of tDCS on motor learning and memory formation: A consensus and critical position paper. Clin. Neurophysiol. 128, 589–603. https://doi.org/10.1016/j.clinph.2017.01.004

Cancel, L.M., Arias, K., Bikson, M., Tarbell, J.M., 2018. Direct current stimulation of endothelial monolayers induces a transient and reversible increase in transport due to the electroosmotic effect. Sci. Rep. 8. https://doi.org/10.1038/s41598-018-27524-9

Cavaliere, C., Aiello, M., Di Perri, C., Amico, E., Martial, C., Thibaut, A., Laureys, S., Soddu, A., 2016. Functional connectivity substrates for tDCS response in minimally conscious state patients. Front. Cell. Neurosci. 10.

Chaieb, L., Kovacs, G., Cziraki, C., Greenlee, M., Paulus, W., Antal, A., 2009. Short-duration transcranial random noise stimulation induces blood oxygenation level dependent response attenuation in the human motor cortex. Exp. Brain Res. 198, 439–444. https://doi.org/10.1007/s00221-009-1938-7

Chew, T., Ho, K.-A., Loo, C.K., 2015. Inter- and intra-individual variability in response to transcranial direct current stimulation (tDCS) at varying current intensities. Brain Stimul. 1–8. https://doi.org/10.1016/j.brs.2015.07.031

Chib, V.S., Yun, K., Takahashi, H., Shimojo, S., 2013. Noninvasive remote activation of the ventral midbrain by transcranial direct current stimulation of prefrontal cortex. Transl. Psychiatry 3, e268.

Clark, V.P., Coffman, B.A., Mayer, A.R., Weisend, M.P., Lane, T.D.R., Calhoun, V.D., Raybourn, E.M., Garcia, C.M., Wassermann, E.M., 2012. TDCS guided using fMRI significantly accelerates learning to identify concealed objects. Neuroimage 59, 117–128.

Colby, K.M., Parkison, C., 1977. Handedness in autistic children. J. Autism Child. Schizophr. 7, 3–9. https://doi.org/10.1007/BF01531110

Corballis, M.C. %J Pl. biology, 2014. Left brain, right brain: facts and fantasies 12, e1001767.

Costafreda, S.G., Chu, C., Ashburner, J., Fu, C.H.Y., 2009a. Prognostic and Diagnostic Potential of the Structural Neuroanatomy of Depression. PLoS One 4, e6353. https://doi.org/10.1371/journal.pone.0006353

Costafreda, S.G., Khanna, A., Mourao-Miranda, J., Fu, C.H.Y., 2009b. Neural correlates of sad faces predict clinical remission to cognitive behavioural therapy in depression. Neuroreport 20, 637–641. https://doi.org/10.1097/WNR.0b013e3283294159

Cummiford, C.M., Nascimento, T.D., Foerster, B.R., Clauw, D.J., Zubieta, J.-K., Harris, R.E., DaSilva, A.F., 2016. Changes in resting state functional connectivity after repetitive transcranial direct current stimulation applied to motor cortex in fibromyalgia patients. Arthritis Res. Ther. 18, 40.

D’Mello, A.M., Turkeltaub, P.E., Stoodley, C.J., 2017. Cerebellar tDCS modulates neural circuits during semantic prediction: A combined tDCS-fMRI study. J. Neurosci. 37, 1604–1613.

DaSilva, A.F., Volz, M.S., Bikson, M., Fregni, F. %J J. of visualized experiments: J., 2011. Electrode positioning and montage in transcranial direct current stimulation.

Datta, A., Baker, J.M., Bikson, M., Fridriksson, J., 2011. Individualized model predicts brain current flow during transcranial direct-current stimulation treatment in responsive stroke patient. Brain Stimul. 4, 169–174. https://doi.org/10.1016/j.brs.2010.11.001

Datta, A., Bansal, V., Diaz, J., Patel, J., Reato, D., Bikson, M. %J B. stimulation, 2009. Gyri-precise head model of transcranial direct current stimulation: improved spatial focality using a ring electrode versus conventional rectangular pad 2, 201–207. e1.

Datta, A., Truong, D., Minhas, P., Parra, L.C., Bikson, M., 2012. Inter-individual variation during transcranial Direct Current Stimulation and normalization of dose using MRI-derived computational models Inter-individual variation during transcranial Direct Current Stimulation. Front. Psychiatry 3, 91. https://doi.org/10.3389/fpsyt.2012.00091

Deng, Z. De, Lisanby, S.H., Peterchev, A. V., 2013. Electric field depth-focality tradeoff in transcranial magnetic stimulation: Simulation comparison of 50 coil designs. Brain Stimul. 6, 1–13. https://doi.org/10.1016/j.brs.2012.02.005

Dmochowski, J.P., Datta, A., Huang, Y., Richardson, J.D., Bikson, M., Fridriksson, J., Parra, L.C. %J N., 2013. Targeted transcranial direct current stimulation for rehabilitation after stroke 75, 12–19.

Drysdale, A.T., Grosenick, L., Downar, J., Dunlop, K., Mansouri, F., Meng, Y., Fetcho, R.N., Zebley, B., Oathes, D.J., Etkin, A., Schatzberg, A.F., Sudheimer, K., Keller, J., Mayberg, H.S., Gunning, F.M., Alexopoulos, G.S., Fox, M.D., Pascual-Leone, A., Voss, H.U., Casey, B., Dubin, M.J., Liston, C., 2017. Resting-state connectivity biomarkers define neurophysiological subtypes of depression. Nat. Med. 23, 28–38. https://doi.org/10.1038/nm.4246

Edwards, D., Cortes, M., Datta, A., Minhas, P., Wassermann, E.M., Bikson, M., 2013. Physiological and modeling evidence for focal transcranial electrical brain stimulation in humans: a basis for high-definition tDCS. Neuroimage 74, 266–75. https://doi.org/10.1016/j.neuroimage.2013.01.042

Esmaeilpour, Z., Shereen, A.D., Ghobadi-Azbari, P., Datta, A., Woods, A.J., Ironside, M., O’Shea, J., Kirk, U., Bikson, M., Ekhtiari, H., 2019. Methodology for tDCS integration with fMRI. Hum. Brain Mapp. https://doi.org/10.1002/hbm.24908

Evans, C., Bachmann, C., Lee, J.S.A., Gregoriou, E., Ward, N., Bestmann, S., 2020. Dose-controlled tDCS reduces electric field intensity variability at a cortical target site. Brain Stimul. 13, 125–136. https://doi.org/10.1016/j.brs.2019.10.004

Fertonani, A., Ferrari, C., Miniussi, C., 2015. What do you feel if I apply transcranial electric stimulation? Safety, sensations and secondary induced effects. Clin. Neurophysiol. 126, 2181–2188. https://doi.org/10.1016/j.clinph.2015.03.015

Fischer, D.B., Fried, P.J., Ruffini, G., Ripolles, O., Salvador, R., Banus, J., Ketchabaw, W.T., Santarnecchi, E., Pascual-Leone, A., Fox, M.D., 2017. Multifocal tDCS targeting the resting state motor network increases cortical excitability beyond traditional tDCS targeting unilateral motor cortex. Neuroimage.

Fonteneau, C., Mondino, M., Arns, M., Baeken, C., Bikson, M., Brunoni, A.R., Burke, M.J., Neuvonen, T., Padberg, F., Pascual-Leone, A., Poulet, E., Ruffini, G., Santarnecchi, E., Sauvaget, A., Schellhorn, K., Suaud-Chagny, M.F., Palm, U., Brunelin, J., 2019. Sham tDCS: A hidden source of variability? Reflections for further blinded, controlled trials. Brain Stimul. 12, 668–673. https://doi.org/10.1016/j.brs.2018.12.977

Frangou, P., Correia, M., Kourtzi, Z., 2018. GABA, not BOLD, reveals dissociable learning-dependent plasticity mechanisms in the human brain. Elife 7. https://doi.org/10.7554/eLife.35854

Fritschi, L., Divitini, M., Talbot-Smith, A., Knuiman, M., 2007. Left-handedness and risk of breast cancer. Br. J. Cancer 97, 686–7. https://doi.org/10.1038/sj.bjc.6603920

Fu, C.H.Y., Mourao-Miranda, J., Costafreda, S.G., Khanna, A., Marquand, A.F., Williams, S.C.R., Brammer, M.J., 2008. Pattern Classification of Sad Facial Processing: Toward the Development of Neurobiological Markers in Depression. Biol. Psychiatry 63, 656–662. https://doi.org/10.1016/j.biopsych.2007.08.020

Garnett, E.O., Den Ouden, D.B., 2015. Validating a sham condition for use in high definition transcranial direct current stimulation. Brain Stimul. 8, 551–554. https://doi.org/10.1016/j.brs.2015.01.399

Geschwind, N., Behan, P., 1982. Left-handedness: association with immune disease, migraine, and developmental learning disorder. Proc. Natl. Acad. Sci. U. S. A. 79, 5097–100.

Gomez-Tames, J., Asai, A., Hirata, A., 2020. Significant group-level hotspots found in deep brain regions during transcranial direct current stimulation (tDCS): A computational analysis of electric fields. Clin. Neurophysiol. 131, 755–765. https://doi.org/10.1016/j.clinph.2019.11.018

Group, F.-N.B.W., 2016. BEST (biomarkers, endpoints, and other tools) resource.

Hordacre, B., Moezzi, B., Goldsworthy, M.R., Rogasch, N.C., Graetz, L.J., Ridding, M.C., 2017. Resting state functional connectivity measures correlate with the response to anodal transcranial direct current stimulation. Eur. J. Neurosci. 45, 837–845. https://dx.doi.org/10.1111/ejn.13508

Hu, J., Li, Y., Yin, Y., Blue, P.R., Yu, H., Zhou, X., 2017. How do self-interest and other-need interact in the brain to determine altruistic behavior? Neuroimage 157, 598–611.

Huang, Y., Liu, A.A., Lafon, B., Friedman, D., Dayan, M., Wang, X., Bikson, M., Doyle, W.K., Devinsky, O., Parra, L.C. %J E., 2017. Measurements and models of electric fields in the in vivo human brain during transcranial electric stimulation 6, e18834.

Jackson, M.P., Rahman, A., Lafon, B., Kronberg, G., Ling, D., Parra, L.C., Bikson, M., 2016. Animal models of transcranial direct current stimulation: Methods and mechanisms. Clin. Neurophysiol. https://doi.org/10.1016/j.clinph.2016.08.016

Jamil, A., Batsikadze, G., Kuo, H.-I., Labruna, L., Hasan, A., Paulus, W., Nitsche, M.A., 2016. Systematic evaluation of the impact of stimulation intensity on neuroplastic after-effects induced by transcranial direct current stimulation. J. Physiol. 00, 1–16. https://doi.org/10.1113/JP272738

Kasahara, K., Tanaka, S., Hanakawa, T., Senoo, A., Honda, M., 2013. Lateralization of activity in the parietal cortex predicts the effectiveness of bilateral transcranial direct current stimulation on performance of a mental calculation task. Neurosci. Lett. 545, 86–90.

Kasuga, S., Matsushika, Y., Kasashima-Shindo, Y., Kamatani, D., Fujiwara, T., Liu, M., Ushiba Brain, J. %J L.A. of B., Cognition, 2015. Transcranial direct current stimulation enhances mu rhythm desynchronization during motor imagery that depends on handedness 20, 453–468.

Kessler, S.K., Minhas, P., Woods, A.J., Rosen, A., Gorman, C., Bikson, M. %J P. one, 2013. Dosage considerations for transcranial direct current stimulation in children: a computational modeling study 8, e76112.

Kessler, S.K., Turkeltaub, P.E., Benson, J.G., Hamilton, R.H., 2012. Differences in the experience of active and sham transcranial direct current stimulation. Brain Stimul. 5, 155–162. https://doi.org/10.1016/j.brs.2011.02.007

Kim, J.H., Kim, D.W., Chang, W.H., Kim, Y.H., Kim, K., Im, C.H., 2014. Inconsistent outcomes of transcranial direct current stimulation may originate from anatomical differences among individuals: Electric field simulation using individual MRI data. Neurosci. Lett. 564, 6–10. https://doi.org/10.1016/j.neulet.2014.01.054

Kriegeskorte, N., Simmons, W.K., Bellgowan, P.S.F., Baker, C.I., 2009. Circular analysis in systems neuroscience: the dangers of double dipping. Nat. Neurosci. 12, 535–540. https://doi.org/10.1038/nn.2303

Kuo, M.-F., Polanía, R., Nitsche, M., 2016. Physiology of Transcranial Direct and Alternating Current Stimulation, in: Transcranial Direct Current Stimulation in Neuropsychiatric Disorders. Springer International Publishing, Cham, pp. 29–46. https://doi.org/10.1007/978-3-319-33967-2_3

Laakso, I., Tanaka, S., Koyama, S., De Santis, V., Hirata, A. %J B. stimulation, 2015. Inter-subject variability in electric fields of motor cortical tDCS 8, 906–913.

Lagopoulos, J., Degabriele, R., 2008. Feeling the heat: the electrode–skin interface during DCS. Acta Neuropsychiatr. 20, 98–100. https://doi.org/10.1111/j.1601-5215.2008.00274.x

Lefaucheur, J.-P., Antal, A., Ayache, S.S., Benninger, D.H., Brunelin, J., Cogiamanian, F., Cotelli, M., De Ridder, D., Ferrucci, R., Langguth, B., Marangolo, P., Mylius, V., Nitsche, M.A., Padberg, F., Palm, U., Poulet, E., Priori, A., Rossi, S., Schecklmann, M., Vanneste, S., Ziemann, U., Garcia-Larrea, L., Paulus, W., 2016. Evidence-based guidelines on the therapeutic use of transcranial direct current stimulation (tDCS). Clin. Neurophysiol. 128, 56–92. https://doi.org/10.1016/j.clinph.2016.10.087

Li, L.M., Uehara, K., Hanakawa, T., 2015. The contribution of interindividual factors to variability of response in transcranial direct current stimulation studies. Front. Cell. Neurosci. 9, 181. https://doi.org/10.3389/fncel.2015.00181

Li, L.M., Violante, I.R., Leech, R., Ross, E., Hampshire, A., Opitz, A., Rothwell, J.C., Carmichael, D.W., Sharp, D.J., 2019. Brain state and polarity dependent modulation of brain networks by transcranial direct current stimulation. Hum. Brain Mapp. 40, 904–915. https://doi.org/10.1002/hbm.24420

Liebetanz, D., Nitsche, M.A., Tergau, F., Paulus, W., 2002. Pharmacological approach to the mechanisms of transcranial DC-stimulation-induced after-effects of human motor cortex excitability. Brain 125, 2238–47. https://doi.org/10.1093/brain/awf238

Ligneul, R., Obeso, I., Ruff, C.C., Dreher, J.-C., 2016. Dynamical Representation of Dominance Relationships in the Human Rostromedial Prefrontal Cortex. Curr. Biol. 26, 3107–3115.

Lin, C.H.J., Chiang, M.C., Wu, A.D., Iacoboni, M., Udompholkul, P., Yazdanshenas, O., Knowlton, B.J., 2012. Age related differences in the neural substrates of motor sequence learning after interleaved and repetitive practice. Neuroimage 62, 2007–2020. https://doi.org/10.1016/j.neuroimage.2012.05.015

Lin, R.L., Douaud, G., Filippini, N., Okell, T.W., Stagg, C.J., Tracey, I., 2017. Structural connectivity variances underlie functional and behavioral changes during pain relief induced by neuromodulation. Sci. Rep. 7.

Loo, C.K., Husain, M.M., McDonald, W.M., Aaronson, S., O’Reardon, J.P., Alonzo, A., Weickert, Cynthia Shannon., Martin, D.M., McClintock, S.M., Mohan, A., Lisanby, S.H., Lisanby, S.H., Krystal, A.D., Peterchev, A. V., McDonald, W.M., O’Reardon, J.P., Aaronson, S., Davis, W., Sklar, J., Loo, C.K., Alonzo, A., Weickert, Cyndi S., Martin, D.M., Mohan, A., Colagiuri, B., Galvez, V., Husain, M.M., McClintock, S.M., 2018. International randomized-controlled trial of transcranial Direct Current Stimulation in depression. Brain Stimul. 11, 125–133. https://doi.org/10.1016/j.brs.2017.10.011

Minhas, P., Bikson, M., Woods, A.J., Rosen, A.R., Kessler, S.K., 2012. Transcranial direct current stimulation in pediatric brain: a computational modeling study. Conf. Proc…. Annu. Int. Conf. IEEE Eng. Med. Biol. Soc. IEEE Eng. Med. Biol. Soc. Annu. Conf. 2012, 859–62. https://doi.org/10.1109/EMBC.2012.6346067

Miranda, P.C., Faria, P., Hallett, M. %J C.N., 2009. What does the ratio of injected current to electrode area tell us about current density in the brain during tDCS? 120, 1183–1187.

Mishima, T., Nagai, T., Yahagi, K., Akther, S., Oe, Y., Monai, H., Kohsaka, S., Hirase, H., 2019. Transcranial Direct Current Stimulation (tDCS) Induces Adrenergic Receptor-Dependent Microglial Morphological Changes in Mice. eNeuro 6. https://doi.org/10.1523/ENEURO.0204-19.2019

Mizuguchi, N., Uehara, S., Hirose, S., Yamamoto, S., Naito, E., 2016. Neuronal Substrates Underlying Performance Variability in Well-Trained Skillful Motor Task in Humans. Neural Plast. 2016.

Moher, D., Liberati, A., Tetzlaff, J., Altman, D.G. %J I. journal of surgery, 2010. Preferred reporting items for systematic reviews and meta-analyses: the PRISMA statement 8, 336–341.

Moisa, M., Polanía, R., Grueschow, M., Ruff, C.C., 2016. Brain Network Mechanisms Underlying Motor Enhancement by Transcranial Entrainment of Gamma Oscillations. J. Neurosci. 36, 12053–12065. https://doi.org/10.1523/JNEUROSCI.2044-16.2016

Monai, H., Hirase, H., 2016. Astrocytic calcium activation in a mouse model of tDCS-Extended discussion. Neurogenes. (Austin, Tex.) 3, e1240055. https://doi.org/10.1080/23262133.2016.1240055

Mondino, M., Ghumman, S., Gane, C., Renauld, E., Whittingstall, K., Fecteau, S., 2019. Effects of Transcranial Stimulation With Direct and Alternating Current on Resting-State Functional Connectivity: An Exploratory Study Simultaneously Combining Stimulation and Multiband Functional Magnetic Resonance Imaging. Front. Hum. Neurosci. 13, 474. https://doi.org/10.3389/fnhum.2019.00474

Monte-Silva, K., Kuo, M.F., Hessenthaler, S., Fresnoza, S., Liebetanz, D., Paulus, W., Nitsche, M.A., 2013. Induction of late LTP-like plasticity in the human motor cortex by repeated non-invasive brain stimulation. Brain Stimul. 6, 424–432. https://doi.org/10.1016/j.brs.2012.04.011

Nasseri, P., Nitsche, M.A., Ekhtiari, H., 2015. A framework for categorizing electrode montages in transcranial direct current stimulation. Front. Hum. Neurosci. 9, 54. https://doi.org/10.3389/fnhum.2015.00054

Nihonsugi, T., Ihara, A., Haruno, M., 2015. Selective increase of intention-based economic decisions by noninvasive brain stimulation to the dorsolateral prefrontal cortex. J. Neurosci. 35, 3412–3419.

Nishida, K., Koshikawa, Y., Morishima, Y., Yoshimura, M., Katsura, K., Ueda, S., Ikeda, S., Ishii, R., Pascual-Marqui, R., Kinoshita, T., 2019. Pre-stimulus Brain Activity Is Associated With State-Anxiety Changes During Single-Session Transcranial Direct Current Stimulation. Front. Hum. Neurosci. 13, 266. https://doi.org/10.3389/fnhum.2019.00266

Nitsche, M.A., Bikson, M., Bestmann, S., 2015. On the use of meta-analysis in neuromodulatory non-invasice brain stimulation. Brain Stimul. https://doi.org/10.1016/j.brs.2015.03.008.This

Nitsche, M.A., Fricke, K., Henschke, U., Schlitterlau, A., Liebetanz, D., Lang, N., Henning, S., Tergau, F., Paulus, W., 2003. Pharmacological modulation of cortical excitability shifts induced by transcranial direct current stimulation in humans. J. Physiol. 553, 293–301.

Nitsche, M.A., Liebetanz, D., Schlitterlau, A., Henschke, U., Fricke, K., Frommann, K., Lang, N., Henning, S., Paulus, W., Tergau, F., 2004. GABAergic modulation of DC stimulation-induced motor cortex excitability shifts in humans. Eur. J. Neurosci. 19, 2720–2726. https://doi.org/10.1111/j.0953-816X.2004.03398.x

Nitsche, M.A., Paulus, W., 2001. Sustained excitability elevations induced by transcranial DC motor cortex stimulation in humans Sustained excitability elevations induced by transcranial DC motor cortex stimulation in. Neurology 57, 1899–1901. https://doi.org/10.1212/WNL.57.10.1899

Nitsche, M.A., Paulus, W., 2000. Excitability changes induced in the human motor cortex by weak transcranial direct current stimulation. J. Physiol. 527 Pt 3, 633–9. https://doi.org/PHY_1055_[pii]

Oldfield, R.C. %J N., 1971. The assessment and analysis of handedness: the Edinburgh inventory 9, 97–113.

Opitz, A., Paulus, W., Will, S., Antunes, A., Thielscher, A., 2015. Determinants of the electric field during transcranial direct current stimulation. Neuroimage 109, 140–150. https://doi.org/10.1016/j.neuroimage.2015.01.033

Palm, U., Reisinger, E., Keeser, D., Kuo, M.-F., Pogarell, O., Leicht, G., Mulert, C., Nitsche, M.A., Padberg, F. %J B. stimulation, 2013. Evaluation of sham transcranial direct current stimulation for randomized, placebo-controlled clinical trials 6, 690–695.

Pascual-Leone, A., Valls-sole, J., Wassermann, E.M., Hallett, M., 1994. Responses to rapid-rate transcranial magnetic stimulation of the human motor cortex. Brain 117, 847–858. https://doi.org/10.1093/brain/117.4.847

Paus, T., 2005. Inferring causality in brain images: a perturbation approach. Philos. Trans. R. Soc. Lond. B. Biol. Sci. 360, 1109–14. https://doi.org/10.1098/rstb.2005.1652

Polanía, R., Nitsche, M.A., Ruff, C.C., 2018. Studying and modifying brain function with non-invasive brain stimulation. Nat. Neurosci. https://doi.org/10.1038/s41593-017-0054-4

Polanía, R., Paulus, W., Nitsche, M.A., 2012. Reorganizing the intrinsic functional architecture of the human primary motor cortex during rest with non-invasive cortical stimulation. PLoS One 7, e30971.

Poreisz, C., Boros, K., Antal, A., Paulus, W., 2007. Safety aspects of transcranial direct current stimulation concerning healthy subjects and patients. Brain Res. Bull. 72, 208–214. https://doi.org/10.1016/j.brainresbull.2007.01.004

Reato, D., Rahman, A., Bikson, M., Parra, L.C., 2013. Effects of weak transcranial alternating current stimulation on brain activity-a review of known mechanisms from animal studies. Front. Hum. Neurosci. 7, 687. https://doi.org/10.3389/fnhum.2013.00687

Ridding, M.C., Ziemann, U., 2010. Determinants of the induction of cortical plasticity by non-invasive brain stimulation in healthy subjects. J. Physiol. 588, 2291–2304. https://doi.org/10.1113/jphysiol.2010.190314

Rosso, C., Perlbarg, V., Valabregue, R., Céline, A., Ferrieux, S., Alshawan, B., Vargas, P., Leger, A., Zavanone, C., Corvol, J.C., Meunier, S., Lehéricy, S., Samson, Y., 2014a. Broca’s area damage is necessary but not sufficient to induce after-effects of cathodal tDCS on the unaffected hemisphere in post-stroke aphasia. Elsevier.

Rosso, C., Valabregue, R., Arbizu, C., Ferrieux, S., Vargas, P., Humbert, F., Attal, Y., Messé, A., Zavanone, C., Meunier, S., Cohen, L., Delmaire, C., Thielscher, A., Herz, D.M., Siebner, H.R., Samson, Y., Lehéricy, S., 2014b. Connectivity between right inferior frontal gyrus and supplementary motor area predicts after-effects of right frontal cathodal tDCS on picture naming speed. Brain Stimul. 7, 122–129. https://doi.org/10.1016/j.brs.2013.08.007

Saiote, C., Polanía, R., Rosenberger, K., Paulus, W., Antal, A., 2013. High-frequency TRNS reduces BOLD activity during visuomotor learning. PLoS One 8, e59669.

Sankarasubramanian, V., Cunningham, D.A., Potter-Baker, K.A., Beall, E.B., Roelle, S.M., Varnerin, N.M., Machado, A.G., Jones, S.E., Lowe, M.J., Plow, E.B., 2017. Transcranial Direct Current Stimulation Targeting Primary Motor Versus Dorsolateral Prefrontal Cortices: Proof-of-Concept Study Investigating Functional Connectivity of Thalamocortical Networks Specific to Sensory-Affective Information Processing. Brain Connect. 7, 182–196.

Saturnino, G.B., Antunes, A., Thielscher, A., 2015. On the importance of electrode parameters for shaping electric field patterns generated by tDCS. Neuroimage 120, 25–35. https://doi.org/10.1016/j.neuroimage.2015.06.067

Schulz, R., Gerloff, C., Hummel, F.C., 2013. Non-invasive brain stimulation in neurological diseases. Neuropharmacology. https://doi.org/10.1016/j.neuropharm.2012.05.016

Simonsmeier, B.A., Grabner, R.H., Hein, J., Krenz, U., Schneider, M., 2018. Electrical brain stimulation (tES) improves learning more than performance: A meta-analysis. Neurosci. Biobehav. Rev. 84, 171–181. https://doi.org/10.1016/j.neubiorev.2017.11.001

Simonsohn, U., Nelson, L.D., Simmons, J.P., 2014. p-Curve and Effect Size: Correcting for Publication Bias Using Only Significant Results. Perspect. Psychol. Sci. 9, 666–681. https://doi.org/10.1177/1745691614553988

Sperling, W., Frank, H., Martus, P., Mader, R., Barocka, A., Walter, H., Lesch, O.M. %J A., Alcoholism, 2000. The concept of abnormal hemispheric organization in addiction research 35, 394–399.

Stagg, C.J., Antal, A., Nitsche, M.A., 2018. Physiology of Transcranial Direct Current Stimulation. J. ECT 1. https://doi.org/10.1097/YCT.0000000000000510

Stagg, C.J., Lin, R.L., Mezue, M., Segerdahl, A., Kong, Y., Xie, J., Tracey, I., 2013. Widespread modulation of cerebral perfusion induced during and after transcranial direct current stimulation applied to the left dorsolateral prefrontal cortex. J. Neurosci. 33, 11425–11431.

Stagg, C.J., Nitsche, M.A., 2011. Physiological Basis of Transcranial Direct Current Stimulation. Neurosci. 17, 37–53.

Tagliazucchi, E., Laufs, H., 2014. Decoding Wakefulness Levels from Typical fMRI Resting-State Data Reveals Reliable Drifts between Wakefulness and Sleep. Neuron 82, 695–708. https://doi.org/10.1016/j.neuron.2014.03.020

Van den Eynde, F., Broadbent, H., Guillaume, S., Claudino, A., Campbell, I.C., Schmidt, U. %J E.P., 2012. Handedness, repetitive transcranial magnetic stimulation and bulimic disorders 27, 290–293.

van Minde, D., Klaming, L., Weda, H., 2013. Pinpointing Moments of High Anxiety During an MRI Examination. Int. J. Behav. Med. 1–9. https://doi.org/10.1007/s12529-013-9339-5

Violante, I.R., Li, L.M., Carmichael, D.W., Lorenz, R., Leech, R., Hampshire, A., Rothwell, J.C., Sharp, D.J., 2017. Externally induced frontoparietal synchronization modulates network dynamics and enhances working memory performance. Elife 6. https://doi.org/10.7554/eLife.22001

Voon, P.J., Kong, H.L., 2011. Tumour genetics and genomics to personalise cancer treatment. Ann. Acad. Med. Singapore 40, 362–8.

Wagner, T., Fregni, F., Fecteau, S., Grodzinsky, A., Zahn, M., Pascual-Leone, A., 2007. Transcranial direct current stimulation: A computer-based human model study. Neuroimage 35, 1113–1124.

Walther, A., Johnstone, E., Swanton, C., Midgley, R., Tomlinson, I., Kerr, D., 2009. Genetic prognostic and predictive markers in colorectal cancer. Nat. Rev. Cancer 9, 489–499. https://doi.org/10.1038/nrc2645

Wang, Y., Ma, N., He, X., Li, N., Wei, Z., Yang, L., Zha, R., Han, L., Li, X., Zhang, D. %J N., 2017. Neural substrates of updating the prediction through prediction error during decision making 157, 1–12.

Wiethoff, S., Hamada, M., Rothwell, J.C. %J B. stimulation, 2014. Variability in response to transcranial direct current stimulation of the motor cortex 7, 468–475.

Woods, A.J., Antal, A., Bikson, M., Boggio, P.S., Brunoni, A.R., Celnik, P., Cohen, L.G., Fregni, F., Herrmann, C.S., Kappenman, E.S., Knotkova, H., Liebetanz, D., Miniussi, C., Miranda, P.C., Paulus, W., Priori, A., Reato, D., Stagg, C., Wenderoth, N., Nitsche, M.A., 2016. A technical guide to tDCS, and related non-invasive brain stimulation tools. Clin. Neurophysiol. 127, 1031–1048. https://doi.org/10.1016/j.clinph.2015.11.012

Woods, A.J., Bryant, V., Sacchetti, D., Gervits, F., Hamilton, R., 2015. Effects of Electrode Drift in Transcranial Direct Current Stimulation. Brain Stimul. 8, 515–9. https://doi.org/10.1016/j.brs.2014.12.007

Woods, A.J., Hamilton, R.H., Kranjec, A., Minhaus, P., Bikson, M., Yu, J., Chatterjee, A., 2014. Space, time, and causality in the human brain. Neuroimage 92, 285–297.

Wu, Q., Chang, C., Xi, S., Huang, I., Liu, Z., Juan, C., Wu, Y., Fan, J., 2015. A critical role of temporoparietal junction in the integration of top‐down and bottom‐up attentional control. Hum. Brain Mapp. 36, 4317–4333.

Xue, G., Juan, C.-H., Chang, C.-F., Lu, Z.-L., Dong, Q., 2012. Lateral prefrontal cortex contributes to maladaptive decisions. Proc. Natl. Acad. Sci. 109, 4401–4406.

Zhang, Y., Yu, H., Yin, Y., Zhou, X., 2016. Intention Modulates the Effect of Punishment Threat in Norm Enforcement via the Lateral Orbitofrontal Cortex. J. Neurosci. 36, 9217–9226.

